# AcceleRest: A Physiology-Aware Masked Autoencoder for Wrist Accelerometer-based Sleep Staging and Apnea Evaluation

**DOI:** 10.64898/2026.01.28.26345056

**Authors:** N.R. Lorenzen, A. Brink-Kjaer, P.J. Jennum, E. Mignot

## Abstract

Sleep is essential for physical and mental health, yet large-scale assessment of sleep stages and sleep apnea is limited by the cost and burden of clinical polysomnography. Wrist accelerometry provides a scalable lower-fidelity alternative, but its usefulness is highly reliant on modeling choices. Generative self-supervised learning has remained unexplored for this purpose. To address this, we developed and pretrained physiology-aware masked autoencoders to capture pulse-and respiration-related motion using ∼700,000 days of wrist accelerometry from 108,904 recordings in the UK Biobank cohort. This effort culminated in AcceleRest, a transformer model pretrained using a respiratory amplification objective. Performance was validated against polysomnography across 478 recordings from six cohorts and devices including independent external test cohorts. AcceleRest feature vectors enabled linear wake–NREM–REM sleep staging with a macro F1 score of 0.69 and respiratory event detection with a macro F1 of 0.56. The combined model outputs enabled sleep apnea severity evaluation with a 67% sensitivity and 96% specificity for severe apnea. Overall agreement between polysomnography and AcceleRest showed a bias of 0.8 min for total sleep duration, with 95% limits of agreement (LoA) of –101.6 to 103.2 min, and 32.5 min for REM sleep duration and 95% LoA of –68.7 to 133.6 min. These findings demonstrate that physiology-aware pretraining can enable robust and clinically meaningful sleep phenotyping from wrist accelerometers, supporting scalable screening and longitudinal monitoring of sleep health. To the best of our knowledge, AcceleRest represents the first wrist accelerometry model for joint sleep stage and apnea evaluation. **All code and models will be made available upon final peer-reviewed publication.**

## Introduction

Sleep is essential for health and well-being^1–4^. The composition of rapid-eye-movement (REM) and non-REM (NREM) sleep stages is important for long-term health outcomes^5^. Sleep apnea represents another major contributor to sleep-related morbidity and is a prevalent^6,7^, underdiagnosed^8^, and treatable sleep disorder associated with severe socioeconomic and health outcomes^9^. The gold-standard polysomnography (PSG) for sleep staging and apnea diagnosis is expensive and burdensome, limiting its use in at-home measurement. Despite developments, there remains an unmet need for simple, low-burden, and commonly available alternatives for sleep staging and sleep apnea screening, for use in large multi-night epidemiological studies and personalized healthcare.

Consumer devices such as smartwatches are increasingly used for personal sleep tracking, but the algorithms and datasets used are often proprietary^10–12^, precluding cross-device testing and limiting reproducibility. Published pipelines for wearable sleep staging generally rely on photoplethysmography-derived pulse oximetry while using accelerometry only for summary activity measures^13–19^. However, several large publicly available cohorts have deployed wrist-worn accelerometers without photoplethysmography, limiting their retrospective applicability^20–22^. Wrist-worn accelerometry devices have been used for recording sleep for more than four decades^23^, but traditional algorithms rely on simple activity counts and primarily distinguish between sleep and wakefulness^23–25^. Hand-crafted features have not substantially improved on legacy algorithms for sleep-wake scoring^26,27^ or enabled sleep staging^28,29^, whereas supervised^30^ and self-supervised^31^ deep learning models have shown promise. However, modern generative pretraining approaches such as transformer masked autoencoders^32^ have not yet been explored for accelerometry-based sleep staging.

High-resolution wrist accelerometers may detect faint motion at the wrist arising from pulse waves and respiratory thoracic expansion during sleep and quiet wakefulness^33–40^. These low-amplitude signals are not captured in the hand-crafted features of previous accelerometry sleep staging studies^26,28^, and are likely missed in previous self-supervised learning approaches^41,42^. Since heart and respiratory rates vary across sleep stages^43,44^, there could be unrealized potential for wrist accelerometry in sleep staging. Apneas also affect these signals^39^, enabling their use in apnea detection using accelerometry and gyroscope recordings^45,46^. However, previous studies have lacked external validation^45–47^.

We aimed to develop a model capable of joint sleep staging and sleep apnea evaluation using wrist accelerometry and to validate its performance across cohorts and devices. We present AcceleRest, a transformer-based model pretrained on ∼700,000 days of accelerometry recordings from the UK Biobank cohort^21^. Building on recent advances in generative self-supervised learning for accelerometry, such as frequency-aware masked autoencoders for downstream activity recognition^41^, we developed pretraining objectives with an explicit focus on frequency ranges associated with pulse and respiratory signals. We propose a respiratory amplification loss, which was used to train the final AcceleRest model.

## Results

Figure 1 shows our model training pipeline. First, the AcceleRest encoder was pretrained on the ∼700,000 days of accelerometry recordings from the UK Biobank. The model inputs consist of 128 minutes of tri-axial accelerometry sampled at 30 Hz, divided into 30 second patches which are masked at random. The respiratory amplification objective then tasks the model with selectively reconstructing and amplifying signal in the frequency range associated with respiration. A detailed description of the respiratory amplification objective can be found in methods and figure 7. Next, linear classification heads were trained on the feature vectors from the resulting AcceleRest encoder and evaluated for downstream sleep staging and respiratory event detection. Lastly, the combined model outputs were used for wrist accelerometry-based sleep apnea severity evaluation.

**Figure 1.**
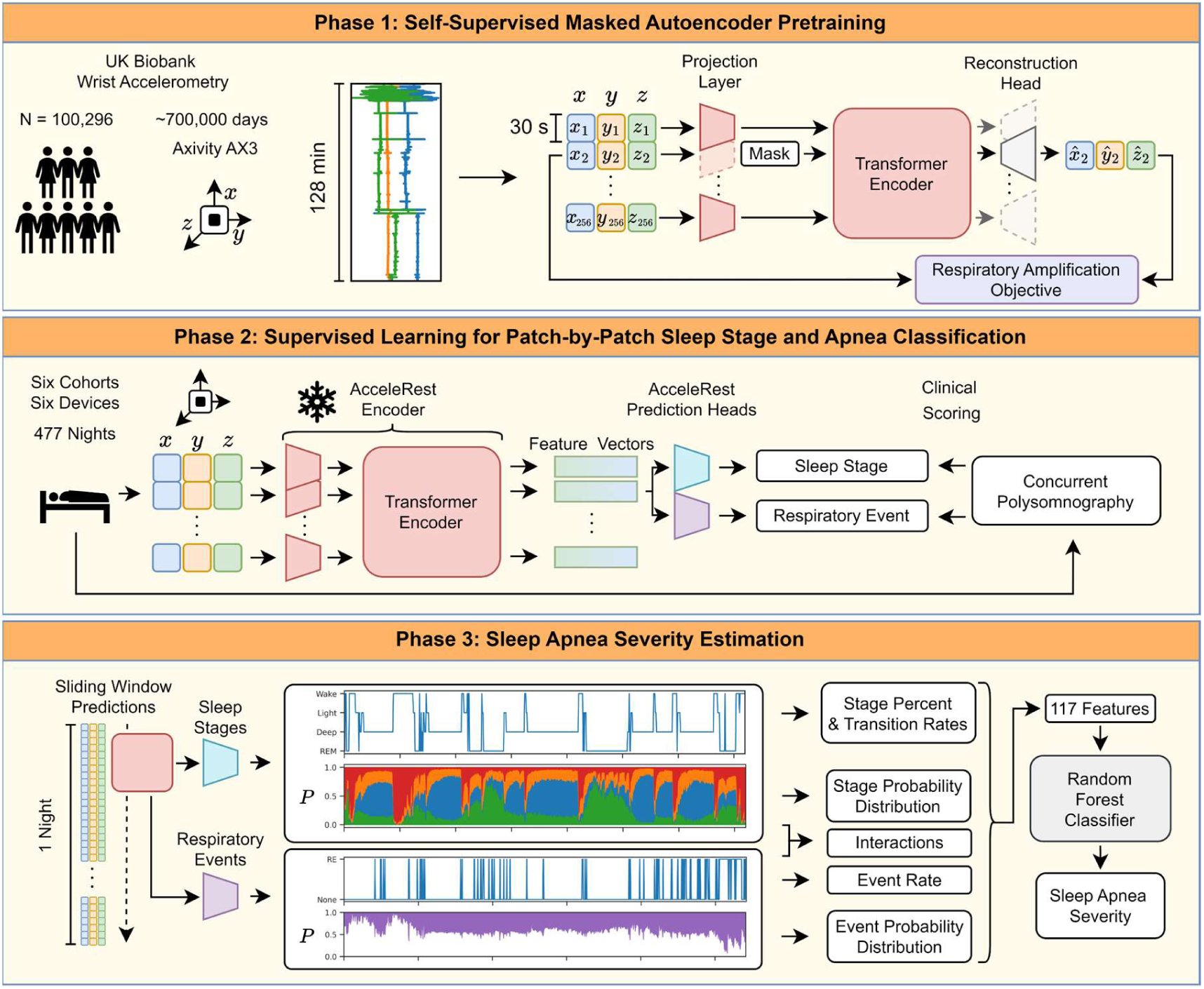
| AcceleRest Development Overview. 1. A masked reconstruction approach focused on amplifying respiratory wrist motions was used to pretrain the AcceleRest encoder on ∼700,000 days of free living tri-axial wrist accelerometry data from the UK biobank. 2. The pretrained AcceleRest encoder was frozen and the feature vectors were used as input to train sleep stage and respiratory event prediction heads against clinical polysomnography annotations. 3. The combined AcceleRest sleep staging and respiratory event detection outputs were used to derive features for a sleep apnea severity classifier.

Table I shows the characteristics of the cohorts used to pretrain and evaluate AcceleRest. Detailed cohort descriptions can be found in methods. Briefly, four cohorts were used to train the classification heads: The traumatic brain injury (TBI)^48^, Dataset for Real-time sleep stage EstimAtion using Multisensor wearable Technology (DREAMT)^49^, the Stanford Technological Analytics and Genomics in Sleep (STAGES)^30^ study, and the Newcastle dataset^50^.

**Table I.** Data.

|  | Pretrain | Train / Internal Test |  |  |  | External Test |  |
| --- | --- | --- | --- | --- | --- | --- | --- |
| Cohort | UKB<sup>27</sup> | TBI | DREAMT | STAGES | Newcastle | Health | SleepAccel |
| Device | Axivity Ax3 | Actiwatch Spectrum | Empatica E4 | Amazfit Arc | GENEActiv | Amazfit Health | Apple Watch |
| N | 115,390 | 260 | 100 | 37 | 28 (55) | 35 | 31 |
| Dropped | 6486 | 22 | 2 | 5 | 0 | 0 | 11 |
| Split | 87:5:8 | 80:20 | 40:60 | 40:60 | 40:60 | 0:100 | 0:100 |
| Age | 62.1±7.9 | 38.4±20.6 | 56.2±16.6 | 38.3±13.6 | 44.9±14.9 | 36.2±13.6 | 29.4±8.5 |
| Sex (% M) | 43.8 | 81.4 | 45 | 45.7 | 61.7 | 40 | 32 |
| BMI | - | 26.2±2.5 | 33.7±8.6 | 29.3±8.5 | - | 28.6±7.8 | - |
| Wake % | - | 24.2 | 25.1 | 27.2 | 30.1 | 28.3 | 7.8 |
| REM % | - | 14.5 | 10.5 | 14.2 | 11.8 | 11.4 | 20 |
| N1 % | - | 9.4 | 11.1 | 5.3 | 4.8 | 2.9 | 6.2 |
| N2 % | - | 31.9 | 49.9 | 41.0 | 35.1 | 41.1 | 51.8 |
| N3 % | - | 19.2 | 3.4 | 12.1 | 18.3 | 15.8 | 12.8 |
| AHI | - | 17.6±20.2 | 22.1±28.7 | 13.1±10.6 | - | 16.0±24.0 | - |
| SA | - | 95/75/36/32 | 35/29/22/13 | 12/14/4/2 | - | 14/15/4/2 | - |

AHI: Apnea-Hypnea Index. SA: Sleep Apnea severity counts (normal / mild / moderate / severe).

These were all divided into training and internal test sets. The Health^30^ and the SleepAccel^18^ cohorts were fully reserved as external test datasets. The Newcastle dataset and SleepAccel cohort did not have apnea annotations available for respiratory event training and apnea severity evaluation. Accelerometry data from these cohorts was recorded using six different devices and encompassed both healthy and disordered sleep.

### Physiology-Aware Self-Supervised vs. Supervised Models

Several objectives were tested before arriving at the respiratory amplification objective as the most effective way to pretrain the AcceleRest encoder. We validated filtering approaches for respiratory and pulse signals that could be integrated into pretraining objectives (Supplementary Tables I and II). Models pretrained with variations on these objectives were then evaluated for downstream sleep staging using linear probing with five-fold cross-validation in the TBI cohort training set.

The best respiration– and pulse-focused encoders were compared to fully supervised models, with or without the extracted pulse and respiration signals as additional input channels (Fig 2a,b). The respiration-focused AcceleRest model performed best overall and the pulse model second best with macro F1 scores of 0.506 ± 0.039 vs. 0.413 ± 0.012 (p = 0.0021, fold-wise paired t-test). The smallest difference between the two pretrained models were for wake with F1 of 0.642 ± 0.033 vs. 0.496 ± 0.032 and the largest was for REM with F1 0.474 ± 0.029 vs. 0.283 ± 0.012.

**Figure 2.**
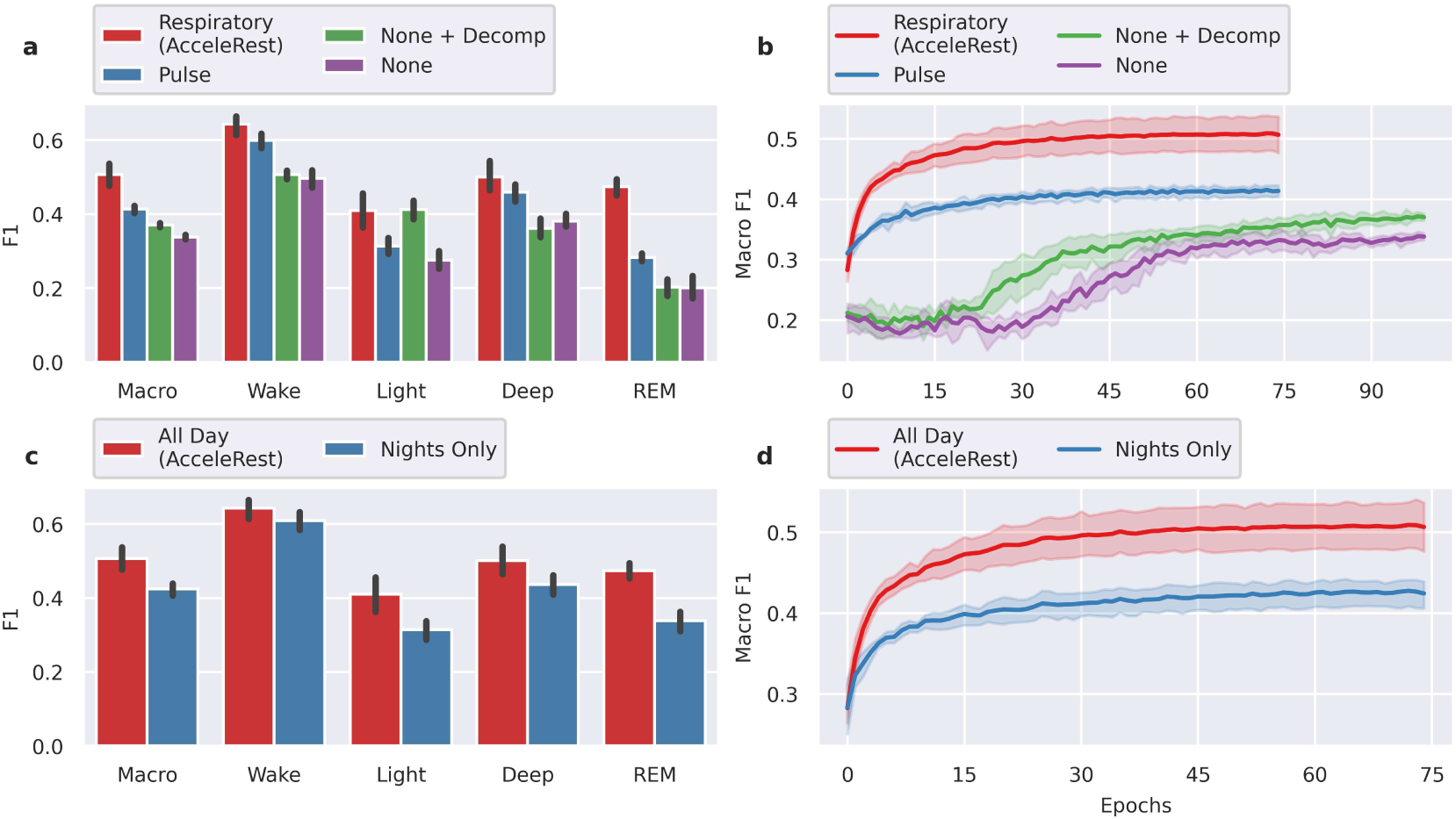
| Model Comparisons. Legend shows model pretraining. **a)** Overall (Macro) and stage-wise F1 scores for linear classifiers trained on feature vectors from respiration– or pulse-focused masked autoencoders and two supervised models on of which included an accelerometry-decomposition layer (None + Decomp) and one without it (None). **b)** Corresponding validation learning curves. **c)** F1 scores for respiration-focused models pretrained using all-day or only nighttime data and **d)** corresponding validation learning curves. Error bars and shaded areas are standard deviations over five cross-validation folds.

Since models were trained with the explicit purpose of downstream sleep scoring, we compared the AcceleRest model, which was trained on all-day data, to a model pretrained with the same respiratory amplification objective only using data from the 12-hour period from 21:00 – 9:00, loosely called nighttime data. We found that training only on nighttime data decreased performance with macro F1 of 0.506 ± 0.039 vs. 0.424 ± 0.021 (p = 0.0014, paired t-test) (Fig. 2c,d). This could be due to decreased heterogeneity in the training data such as less periods of wakeful rest. However, exclusion of patches with SD > 0.015 in the reconstruction loss meant that for the full day data the models are effectively only trained on an average of 36% of the patches, while this number was 70% for the night period data. This suggests that only a very small amount of the additional daytime data is used.

The same experiments were done using a long short-term memory (LSTM) classification head to test the effects of sequence learning with labeled data. Generally, the linear classifier had lower bias but slightly worse classification performance. See supplementary figures 4-6 for comparative results of the classification heads.

### Sleep Scoring Performance

After obtaining the final AcceleRest encoder, the output feature vectors were used to train a linear classification head. This was done in three steps. First, the classifier was trained on the 191 recordings from the TBI cohort training set and evaluated on the test sets of the other internal cohorts. Next, the classifier was trained on the internal training sets and evaluated on the internal test sets. Lastly, the classifier was trained on the internal training and test sets and evaluated on the external test sets.

The subject-wise performance metrics can be found in supplementary tables 5-7. For sleep-wake scoring on the external test set the model achieved a macro F1 score of 0.783 ± 0.116 and Cohen’s kappa of 0.575 ± 0.215. For the three-class wake-NREM-REM scoring it obtained an F1 of 0.648 ± 0.148 and Cohen’s kappa of 0.509 ± 0.214. For the four-class wake-light-deep-REM task, the performance was an F1 of 0.468 ± 0.119 and Cohen’s kappa 0.315 ± 0.147. The values were generally slightly lower on the internal test sets, with F1 of 0.707 ± 0.128 and Cohen’s kappa of 0.437 ± 0.224 for sleep-wake and F1 of 0.541 ± 0.132 and Cohen’s kappa of 0.358 ± 0.189 for wake-NREM-REM. This could be due to a mix of higher difficulty data and less training data available for the internal compared to the external evaluation. In either case these results underscore the generalizability of the AcceleRest feature vectors.

The AcceleRest external test set scores were compared those reported for SleepNet (Supplementary Table 5), a state-of-the-art self-supervised model for wrist accelerometry-based sleep staging^31^. AcceleRest obtained an increase in average subject-wise macro F1 score of 15.7% for wake-NREM-REM and 12.5% for sleep-wake. However, internal test scores were lower for AcceleRest, suggesting that AcceleRest is less susceptible to overfitting compared to SleepNet (Supplementary Table 7). AcceleRest also achieved slightly higher held-out performance even when only trained on the TBI training set and evaluating on the other *unseen* internal cohorts with an increase in macro F1 of +2% compared to the SleepNet external test performance (Supplementary Table 6).

Figure 3 shows the epoch-by-epoch classification results from the internal and external evaluations. In agreement with the subject-wise results the overall F1 score for wake-NREM-REM increased by 11% in the external evaluation (Fig. 4c,f). However, a strong bias for classification of light as deep sleep appeared in the external evaluation, which was not there in the internal evaluation (Fig. 4b,e). Meanwhile, the receiver operating characteristics (ROC) curves revealed largely unchanged area under the curve (AUC) for light and deep sleep. This suggests a shift in label distribution as the reason for the bias, possibly due to the relatively low amount of deep (N3) sleep annotated in the DREAMT cohort (Table 1).

**Figure 3.**
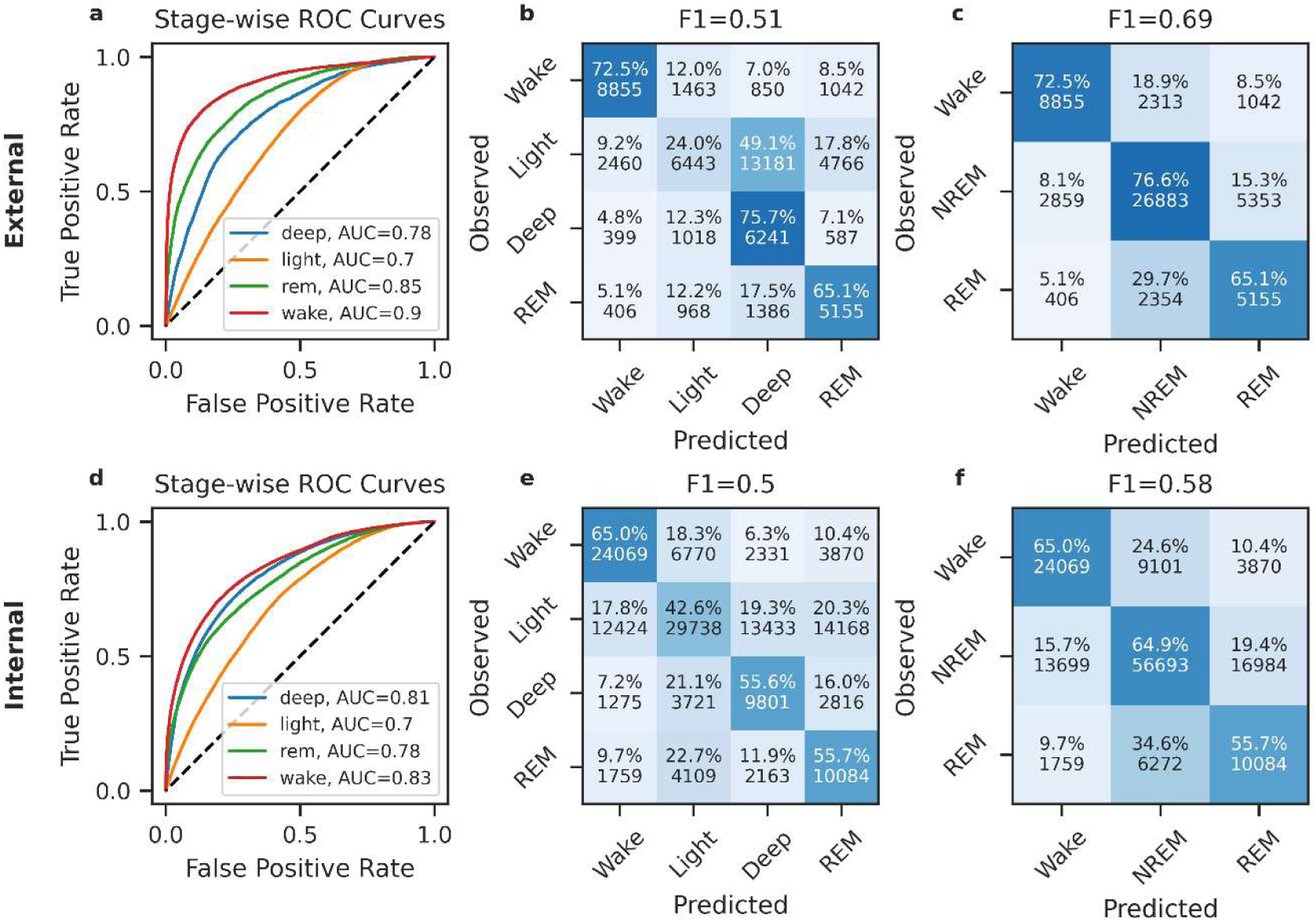
| Epoch-by Epoch Classification Performance. **a,d** Receiver operating characteristic (ROC) curves computed for each sleep stage in a ‘one-vs-rest’ fashion. **b,e)** Four-stage confusion matrices with row-by-row percentages and absolute numbers. **c,f)** Three-stage confusion matrices.

**Figure 4.**
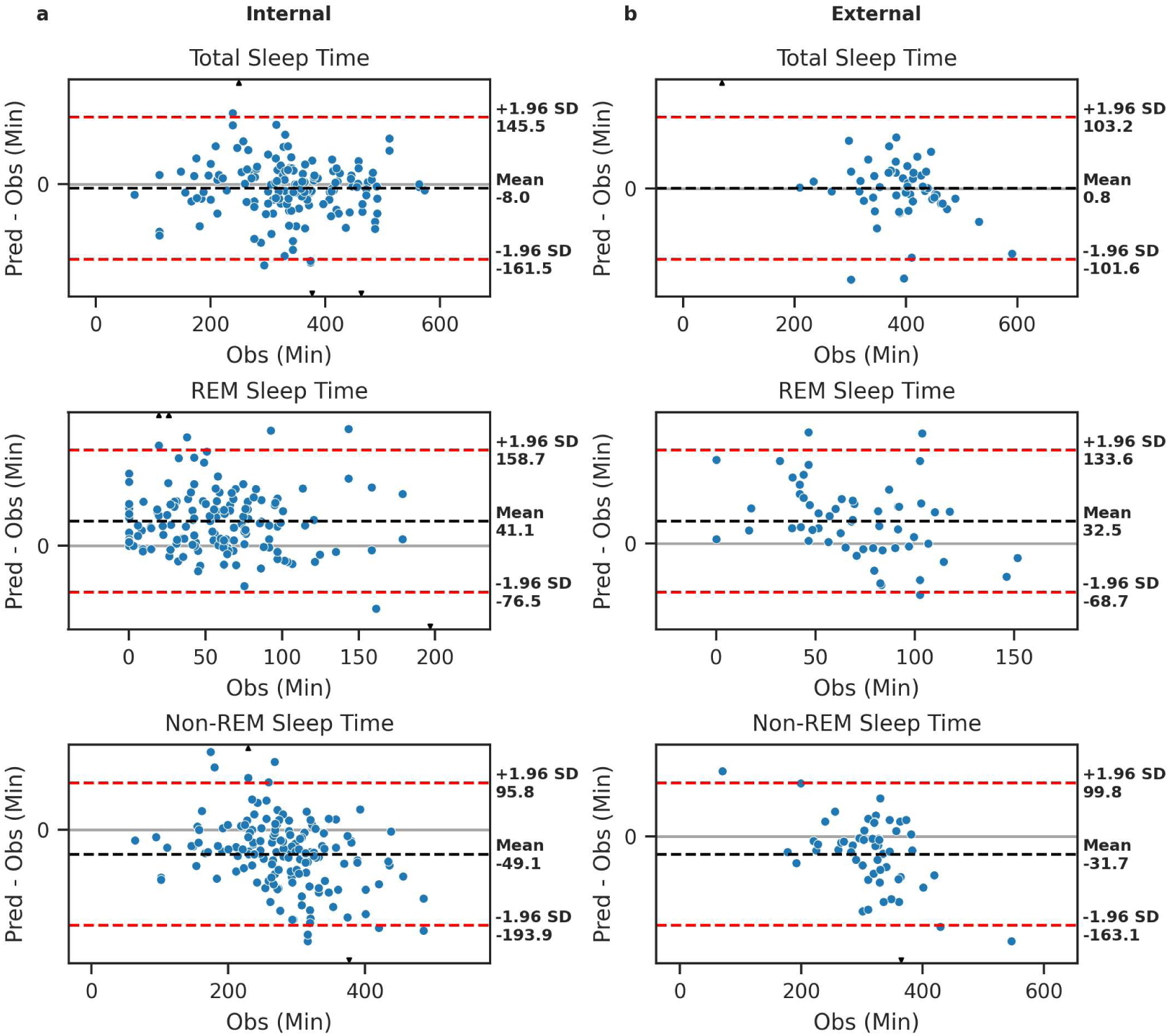
| Summary Measure Residual Plots Agianst Polysomnography. **a)** Total Sleep Time. **b)** Sleep Efficiency. **c)** Total Rapid Eye Movement (REM) sleep. **d)** Total Non-REM sleep. **e)** Total deep sleep. **f)** Total light sleep. g) Sleep Onset Latency. **h)** Wake after sleep onset.

The agreement between PSG and AcceleRest on key summary metrics was quantified using Bland-Altman analysis in the internal and external test sets (Fig. 4). The overall bias for total sleep time was –8 minutes with 95% limits of agreement (LoA) of –161.5 to 145.5 minutes in the internal evaluation and 0.8 minutes and LoA of –101.6 to 103.2 minutes in the external evaluations. AcceleRest tended to overestimate time spent in REM sleep and underestimate time spent in NREM sleep. For REM the bias was 41.1 minutes and LoA –76.5 to 158.7 minutes in the internal and 32.5 minutes with LoA –68.7 to 133.6 in the external evaluations. For NREM the bias was –49.1 minutes and LoA –193.9 to 95.8 minutes in the internal and –31.7 minutes with LoA –163.1 to 99.8 in the external evaluations. The same analysis can be found for an exhaustive list of summary measures in supplementary tables 3 and 4 and the Bland-Altman plots in supplementary figures 9-12.

#### Sleep Apnea Severity Evaluation

First, a linear classification head was trained on the AcceleRest feature vectors to detect the occurrence of apneas and hypopneas, grouped into respiratory events, within each 30-second patch. The respiratory event detection head was trained on the TBI, DREAMT and STAGES cohorts using only the same training partitions used for sleep staging. The model obtained a moderate performance on the external Health dataset with a patch-by-patch macro F1 of 0.56 but with a strong over-estimation of respiratory events (Fig. 5). With external test set subject-wise precision of 0.965 ± 0.051 and a recall of 0.776 ± 0.137 for predicting no event and a precision of 0.131 ± 0.146 and a recall of 0.489 ± 0.265 for respiratory events (Supplementary Table 9). The rate of detected events showed only moderate correlation with the observed apnea-hypopnea index with a Pearson’s R of ∼0.55 (Supplementary Fig. 14).

**Figure 5.**
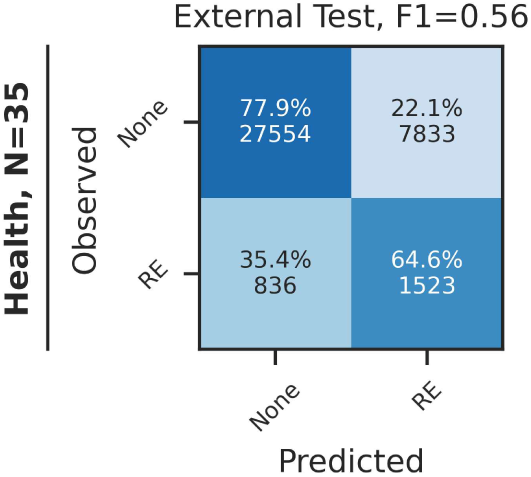
| Confusion Matrix for External Test Set Respiratory Event Detection. The patch-by-patch confusion between classification of no event (None) or an unspecified respiratory event (RE) including central and obstructive apneas and hypopneas.

To more accurately estimate sleep apnea severity, a random forest classifier was trained for sleep apnea severity classification. This was done using 117 features derived from a full night of AcceleRest respiratory event detection and sleep staging outputs. A combination of respiratory event detection, sleep staging, and their temporal interactions were found to be most effective (Supplementary Fig. 17). The internal test partitions were further subdivided into validation and test partitions, with the validation partition used for hyper-parameter optimization. The final validation performance was a macro F1 score of 0.63 ± 0.06 (mean ± standard deviation over 5 cross-validation folds) (Fig. 6a). Finally, the apnea severity classifier was trained on the AcceleRest outputs from the internal training and validation partitions and evaluated both internally on the remaining internal test sets and externally on the Health dataset (Fig. 6b). This was done to increase the number of severe cases available for testing. The combined test performance was lower compared to the internal cross validation with an F1 score of 0.52.

**Figure 6.**
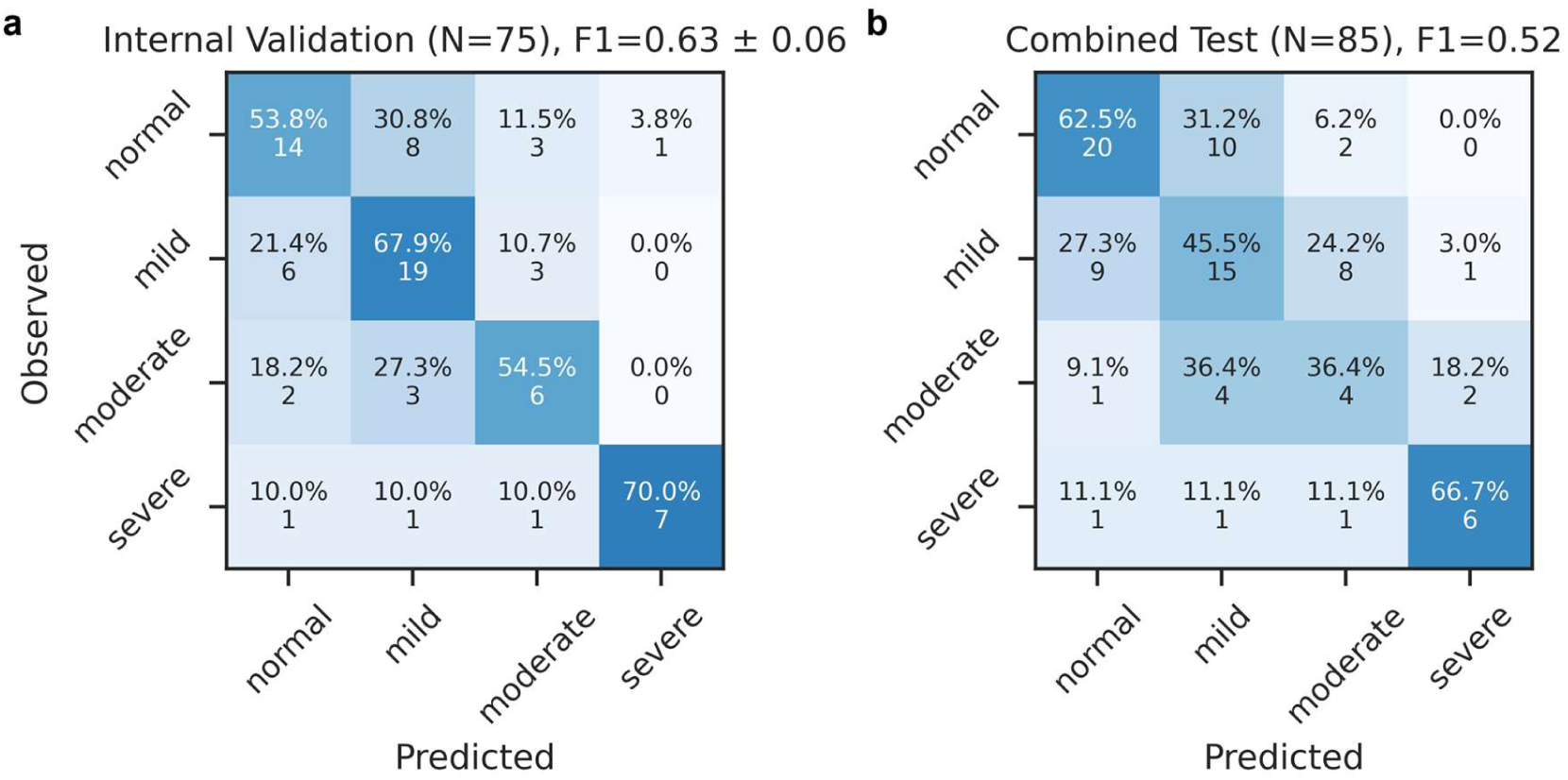
| Validation and Test Set Performance of Sleep Apnea Severity Classifier. **a)** Shows confusion on the internal validation set, with F1 = 0.63 ± 0.06 (mean ± SD) across five validation folds. **b)** Confusion on the combined internal and external test sets.

**Figure 7.**
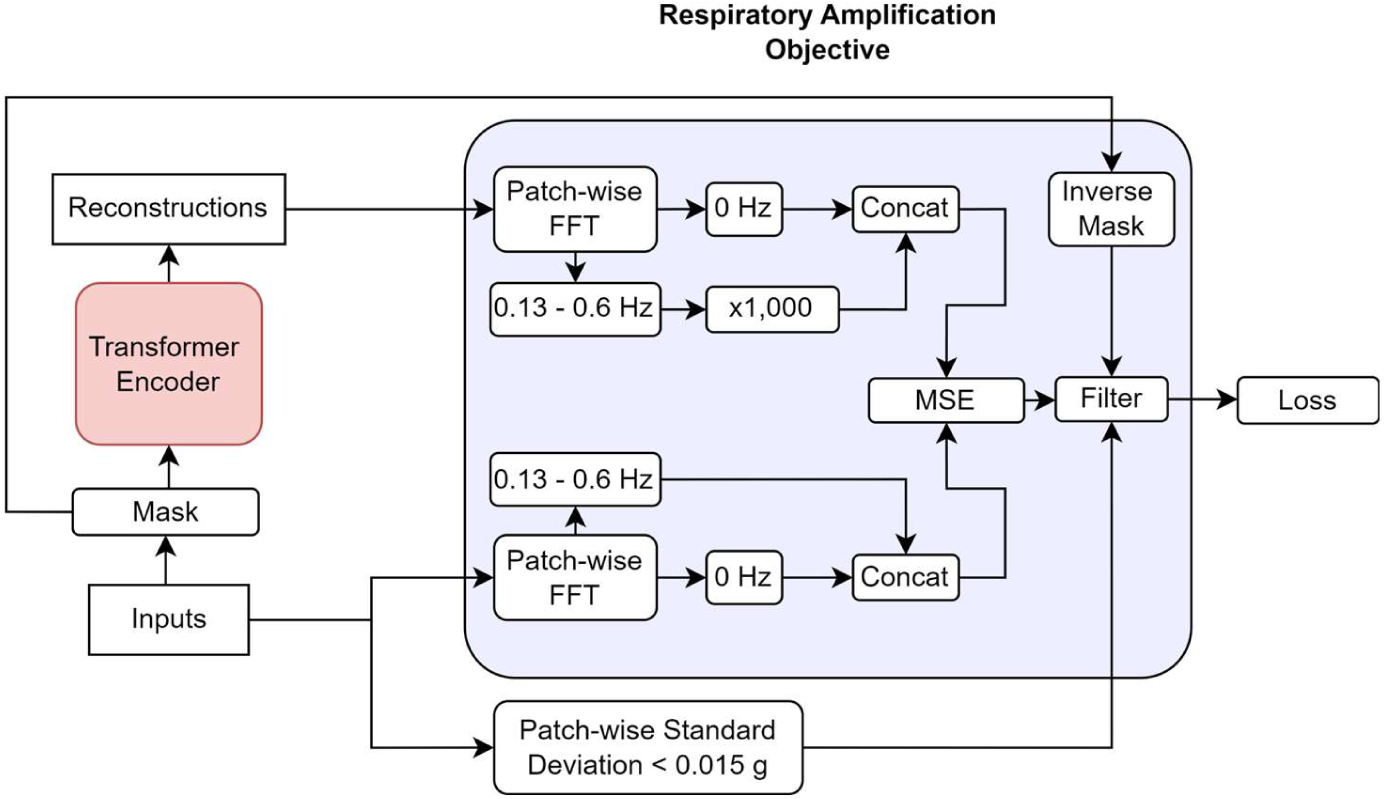
| Respiratory Amplification Objective. The patched input accelerometry signal is masked and reconstructed by the transformer encoder. The Fast Fourier Transform (FFT) is applied patch-wise and to both inputs and reconstructions and frequency magnitudes are extracted. The bins in the 0.13-0.6 Hz frequency range associated with breathing are isolated and selectively amplified by a factor 1000 for the reconstructions, while the 0 Hz bin is kept as reference. Remaining frequency bins are discarded. The mean-squared-error (MSE) loss is calculated between the frequency magnitudes. Only the masked patches with standard deviations < 0.015 are kept in the final loss.

On the combined test set the model correctly classified 6/9 severe cases as severe giving a recall/sensitivity of 67%, 6/9 severe predictions were severe giving a precision/positive predictive value of 67%, and 73/76 non-severe cases were correctly classified as non-severe for a specificity of 96%. For a moderate/severe class the corresponding numbers were a recall of 13/20 = 65%, a precision of 13/24 = 54%, and a specificity of 54/65 = 0.83%. There was no indication that including internal test set inflated the overall performance with an F1 of 0.47 for the internal and 0.48 for the external test sets (Supplementary Fig. 18). The epoch-by-epoch RE detection performance was also comparable on the internal (Supplementary Fig. 13b) and external test sets (Fig. 5). This suggests that the severity estimation performance gap is due to hyper parameter overfitting of the random forest classifier. Feature importances for the apnea severity classification are shown in Supplementary Fig. 18.

## Discussion

In this study, we introduced AcceleRest, a transformer model pretrained specifically for wrist accelerometry-based sleep analysis using ∼700,000 days of data from 108,904 recordings. Our work makes several contributions. First, the general concept of explicitly physiology-aware pretraining for accelerometry. This effectively pushes the boundaries for accelerometry-based sleep analysis by engineering the pretraining task to align better with the downstream goal. The approach not only leverages advances in generative self-supervised learning, but the combination with respiratory signal extraction rather than a signal-agnostic generative approach was key. This is likely due to the small amplitudes of these signal components. The improved performance over supervised models provided with the accelerometry signal decomposed into the respiratory and pulse signals further underlines that the pretraining phase is also essential for model performance.

The AcceleRest feature vectors provided representations that generalized across cohorts and devices and enabled strong linear separability of wakefulness, NREM, and REM sleep (F1 = 0.69), and moderate respiratory event detection performance (F1 = 0.56) in fully held-out cohorts. Sleep staging performance was validated against polysomnography-derived annotations across six cohorts and devices and while apnea evaluation was across four cohorts. The AcceleRest feature vectors were especially effective for the separation of REM sleep compared to models trained with a focus on pulse-related motion and fully supervised models.

The masked autoencoder-style pretraining seemingly does more than simply attune the model to the respiratory signal, possibly by also instilling an inherent sequence understanding of how these signals evolve over time. This is supported by AcceleRest achieving state-of-the-art performance with a linear classifier, contrasting with prior work such as SleepNet which leveraged more a complex supervised sequence model for sleep staging on top of the pretrained single-patch encoder^31^. Furthermore, training a lightweight supervised sequence classifier on the AcceleRest feature vectors resulted in a modest gain of 1% in F1 score for sleep-wake and wake-NREM-REM classification. In combination, this suggests that relevant context information is already encoded in the patch-wise AcceleRest feature vectors. This aligns well with findings of reduced gain from supervised sequence modeling when using masked autoencoders for activity recognition^41^. We also found that despite the slightly increased classification performance, using a sequence classifier also resulted in stronger biases. This is likely due to the learning of statistical patterns in ultradian sleep cycle. Such inductive biases can improve overall classification performance but have detrimental effects on performance for individuals with atypical sleep patterns. Since this is first work to report robust linear classifier performance for accelerometry-based sleep staging, these discrepancies have not previously been investigated.

Performance comparison based on reported metrics is always problematic due to possible distribution shifts between cohorts. This was mitigated by also training the AcceleRest classifier only using the TBI cohort and comparing the performance by using the unseen internal cohorts as an additional external test set. Even in this setting AcceleRest performed slightly better on both sleep-wake and wake-NREM-REM. This is in spite SleepNet being trained on a substantially larger labeled dataset with 1113 recordings^31^, compared to 191 for the single-cohort training. Along with lower internal test scores compared to SleepNet, this shows that AcceleRest relies less on labeled data in the supervised learning phase by providing generalizable feature vectors with stronger linear separability of sleep stages.

The respiratory event detection was not accurate enough to allow direct end-to-end estimation of the apnea-hypopnea index with sufficient accuracy. Previous studies have reported higher accelerometry-based respiratory event detection performance^45–47^. However, none of these have reported performance in an external test set, leaving their generalizability in question. This is the first study to employ a pretrained model for this purpose, which enabled leveraging the dual sleep staging and respiratory event detection capabilities of AcceleRest. Doing this we were able to achieve a high specificity of 96% for severe sleep apnea. With a moderate sensitivity of 67% this approach is not diagnostic but has potential to flag high-risk individuals. We were not able to test the effect of using multiple nights of accelerometry recording in this study, but it is likely that this would further increase the power. Future studies should test the use of multi-night accelerometry recording against PSG. To the best of our knowledge these are the first cross-cohort and device test set results reported for accelerometry-based sleep apnea severity estimation and can thus serve as a benchmark for future studies.

While the respiratory amplification objective enabled the development of our most successful model, this study does not conclude that respiratory modeling is necessarily more effective than pulse modeling. Future studies should investigate modifying the pulse-focused objective or even combining the two, which led to failed model convergence in our attempts. The only evidence that respiration might be preferrable to pulse modelling is our finding that respiration rate is more often accurately determined compared to heart rate. However, previous studies have concluded the opposite^39^. This could be due to differences in the pulse signal extraction pipelines.

Self-supervised learning for accelerometry and especially for downstream sleep analysis is a nascent field, and we assume that the upper limits of what is possible in accelerometry-based sleep staging have yet to be identified. Here, we introduced the concept of physiology-aware masked-autoencoders, demonstrating that domain knowledge integration in pretraining objective design can unlock clinically relevant feature extraction. AcceleRest thus provides a foundation for scalable, low-burden sleep phenotyping in both research and personalized healthcare.

## Methods

### Data and Preprocessing

Tri-axial wrist accelerometry data was resampled to 30 Hz after applying an anti-aliasing filter, and calibrated for gravity using the algorithm described in^51^.

#### UK Biobank

The UK Biobank accelerometry subcohort^21^ was used for model pretraining. It consists of 115,390 recordings of ∼7 days of in-the-wild wrist-worn accelerometry from 103,618 participants. Tri-axial accelerometry was recorded at 100 Hz using Axivity AX3 devices. Recordings for which calibration failed or for which less than 24 hours of data were available were dropped, leaving 108,904 total recordings. These were split subject-wise (including multiple recordings) into 94,876 (87%) training-set recordings from 86,618 subjects, 5,393 (5%) validation set recordings from 4,838 subjects, and 8,635 (8%) test set recordings from 6476 subjects. The validation set recordings were only used to monitor pretraining convergence and the test set is reserved for future work.

#### The Traumatic Brain Injury (TBI) cohort

Tri-axial acceleration data were recorded with a Actiwatch Spectrum. Concurrent night PSG and wrist accelerometry was collected for 271 patients as described in^48^. Out of 260 patients with PSG, wrist accelerometry, and hypnograms, 19 were dropped for having < 2 hours of data, and 3 were excluded due to failed calibration, leaving 238 subjects. The TBI cohort was split 80:20 (N = 191 and 52) for training and testing. The TBI training set was used in five-fold cross validation during model ablations.

#### DREAMT: Dataset for Real-time sleep stage EstimAtion using Multisensor wearable Technology

The DREAMT study includes 100 participants with concurrent nocturnal PSG and wrist accelerometry from Empatica E4 wristband (Empatica Inc., Milano, Italy) on the left wrist. Participants were recruited from the Duke University Health System (DUHS) Sleep Disorder Lab under IRB approval (#Pro00108961) see^49^ for details. Here 2 participants were dropped due to calibration issues. We identified a clock drift between the accelerometry and PSG signal of 15ms/s across all recordings which was corrected by resampling the PSG and hypnogram. The DREAMT cohort was split 40:60 for training and testing (N = 39 and 59).

#### The Stanford Technological Analytics and Genomics in Sleep (STAGES) study

Tri-axial acceleration data was collected using an Amazfit Arc device at 25 Hz sampling rate with 12-bit resolution at a dynamic range of ±8g. Data collection and exclusions for the STAGES actigraphy sub-cohort is described in^30^. Out of 323 recruited participants, data from 201 were lost, and 37 participants with concurrent PSG and wrist accelerometry remained after exclusions. Additionally, 5 participants were excluded due to failed calibration with 32 recordings remaining. These were split 40:60 for training and testing (N = 13 and 19).

#### Health cohort

Tri-axial acceleration was recorded using Amazfit Health (Huami, Inc.) 25 Hz sampling rate with 12-bit resolution at a dynamic range of ±8g. Data collection and exclusions are described in^30^. Out of 54 recruited participants 35 recordings remained after exclusions. These were all used for external testing.

#### SleepAccel

We used the SleepAccel dataset described in^18^. This dataset included 39 participants of which 4 were excluded due to transmission errors, 3 due to sleep apnea, and 1 due to RBD, for a remaining total of 31 participants. Participants wore an Apple Watch (series 2 or 3) concurrently with a night PSG and 7 – 14 days prior. Here, we only used the data with concurrent PSG-derived hypnogram labels. The data had variable sampling rates with most participants having median instantaneous sampling rates of ∼ 50 or 65 Hz. For preprocessing, we first identified sampling gaps of > 1 second and then kept contiguous segments > 4 hours with no such gaps. The resulting segments were uniformly resampled to the median sampling rate using linear interpolation, then filtered for anti-aliasing and resampled to 30 Hz as with all other data. We dropped 7 participants due to having no 4 hours gap-free segments, one participant was dropped due to having no hypnogram labels in the resulting window, and two participants were dropped due to having sampling rates of 10 Hz. This left 21 participants with 4 – 8 hours of wrist accelerometry data with hypnogram labels. All of these were used as an external test set.

#### Newcastle Dataset

The Newcastle Dataset^50^ consists of 55 wrist accelerometry recordings from 28 subject-nights with 28 left-hand recordings and 27 right-hand recordings. Tri-axial accelerometry was recorded with a GENEActiv device at 85.7 Hz. The Newcastle Dataset is a clinical cohort with 21 out of 28 patients having some diagnosis including apnea (n = 8), restless legs syndrome (n = 5), REM sleep behavior disorder (n = 3), narcolepsy (n = 1), insomnia (n = 2), and hypersomnia (n = 2).

### Extraction of physiological signals from wrist accelerometry

For respiratory signal extraction, a 0.13 – 0.6 Hz Fast Fourier Transform (FFT) band-pass filter was applied. For pulse signal extraction we applied an approach similar to those described in^34,37^. First, a 4-14 Hz FFT band-pass filter, was applied. Then, cross-axial vector magnitudes 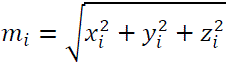, were calculated. Lastly, a pulse-band (0.6 – 1.7 Hz) FFT band-pass filter was applied. The pulse-band filter applied here is narrower than the 0.5 – 3.5 Hz range that is often applied^33,34,37^. The 0.6 – 1.7 Hz range covers heartrates of 36 – 102 beats per minute (BPM) and was found to include the peaks of interest in a spectral analysis (Supplementary Fig. 1). This range does not include all pathologically possible resting heart rates but was chosen to limit the number of higher harmonics and noisy frequency bins models would have to account for during inference. For respiratory and pulse spectrograms, the final inverse FFT was simply skipped. These pipelines were implemented in PyTorch for GPU support and integration with model training. Spectrograms were calculated in 30-second, non-overlapping patches.

### Model Architecture

The models introduced in this paper are encoder-only transformers^52^ with rotational positional encodings^53^. The models take 128 minutes of tri-axial accelerometry sampled at 30 Hz as input. Each non-overlapping 30-second patch is projected by a linear layer to a 256-dimensional embedding space for a sequence of 256 embeddings, which are fed to the transformer module, consisting of 12 transformer blocks. Each transformer block consists of a multi-head self-attention layer with 8 attention heads followed by a feed-forward layer with SwiGLU activations^54^ with a hidden dimension of twice the embedding dimensionality. Root-mean-square layer normalization^55^ is applied in a pre-norm fashion^56^. Finally, a linear layer outputs time-domain reconstructions during pretraining or a classification head outputs patch-wise classifications during down-stream supervised training and inference. Note that either patch or the clinical term epoch, used in sleep scoring, will be used to refer to 30-second patches, not to be confused with model training epochs.

### Pretraining

Models were pretrained for 20 epochs on the UK Biobank accelerometry dataset, in each epoch a random 24-hour period of continuous wear time is sampled from a full multi-day recording. Models were pretrained using masked autoencoder signal reconstruction with the loss applied on frequency magnitudes, similar to what was done in previous work for human activity recognition^57^. In short, 60% of input patch embeddings were replaced with a mask token, and the model inferred a time-domain signal reconstruction for each 30-second patch. FFT magnitudes were computed for the patch-wise reconstructions and the corresponding input patches. This produces 451-bin magnitude spectrograms with a frequency resolution of 0.033 Hz or ∼2 BPM. Either a respiratory or pulse amplification objective was then applied. Each of the objectives are only applied to masked patches with standard deviation < 0.015 g, as higher activity patches are unlikely to contain meaningful signal for respiration and pulse signal extraction^39^.

#### Respiratory Amplification Objective

The model outputs a signal for each input axis and the FFT magnitude spectrum is computed for all three axes. Then the 0 Hz and 0.13 – 0.6 Hz bands are kept and the 0.13 – 0.6 Hz band is amplified 1000-fold for the input patches only. This way the model must reconstruct a time-domain signal that is amplified in the respiratory frequency band relative to the 0 Hz bin. No loss is applied to the remaining frequency bins. In pilot experiments we found that training the model to effectively band-stop filter frequencies outside the 0 Hz and respiratory bands led to poorer convergence. We also found that the full 2 BPM frequency resolution was key for down-stream performance. Using 10 s patches or 30 s patches with 3x frequency domain down-sampling, both resulting in 6 BPM resolution, led to similar poor performance.

#### Pulse Amplification Objective

The multi-step filtering approach involved in pulse signal extraction means that there are multiple possible entry points for the loss objective. The model could either reconstruct the initial 4-14 Hz band-pass filtered signal or reconstruct the final pulse signal directly. The final pulse-focused model was tasked with reconstructing the pulse signal directly with a 10k-fold amplification applied to the target spectrum. For relative amplification both 0 Hz and the 0.13 – 0.6 Hz bands were tested as references as power remains in these bands before the final SCG band-pass filter (Supplementary Fig. 1). However, both these strategies led to poor convergence, and the final pulse model was pretrained without relative amplification. Contrary to the respiratory amplification, the pulse spectra were downsampled to halve the frequency resolution to 4 BPM as this was found to improve convergence.

### Sleep Staging and Respiratory Event Detection

Classifiers for all down-stream tasks were trained on the embeddings of the frozen AcceleRest encoder, except in pretrained model comparisons during ablation studies where another pretrained encoder and fully supervised models were also tested. Classifiers were trained using the inverse-class frequency weighted cross-entropy loss.

Sleep stage and apnea labels were derived by expert scoring according to the American Academy of Sleep Medicine (AASM) criteria^58^. Clinical sleep scoring classifies 30-second patches into wakefulness, REM, and NREM stages 1-3. For model training NREM stage 1 and 2 were combined to light sleep, and N3 was renamed deep sleep. For three-class classification evaluation the light and deep sleep outputs were combined into NREM for wake-NREM-REM, and for two-class sleep-wake NREM and REM evaluation outputs were combined into sleep.

Respiratory event annotations included hypopneas, obstructive-, and central apneas. To obtain patch-wise event labels, any patch which included part of a respiratory event was labeled positive for a respiratory event with priority given to a combined obstructive– and central apnea class.

We tested both combined two-class respiratory event detection and three-class hypopnea and apnea event classification. Supplementary figure 13 shows the event detection confusion matrices and supplementary table 8 shows the combined respiratory event detection performance for two-class and three-class training. The two-class training provided the best overall respiratory event rate estimate shown by the correlation with the observed apnea-hypopnea index (Supplementary Fig. 14). Two-class respiratory event detection also provided the best overall apnea severity estimates in the internal validation (Supplementary Fig. 15). We also tested apnea-hypopnea index regression and found that this consistently led to strong overestimation in the mild severity range (Supplementary Fig. 16).

#### Performance Evaluation

We evaluated both patch-by-patch model performance and subject-wise performance evaluation. In subject-wise evaluation we used a sliding context window with one-epoch steps and averaged the output scores for each class from each context window (See supplementary figures 24-26 for illustration). This means that the number of predictions averaged for each epoch increases from one, for edge-epochs, to 256, effectively using sliding context windows as a form of ensemble prediction. All context window outputs are computed in parallel in a single forward-pass.

As performance metrics, we report precision, recall, F1 score, Cohen’s kappa, accuracy, and Area Under the Receiver operating Curve (AUROC). Overall precision, recall, F1 score metrics are macro-averaged i.e., the unweighted average across classes. For sleep apnea severity evaluation, we also report model specificity.

### Pretrained and Supervised Model Comparisons

In these experiments, the TBI training set consisting of 191 subjects, was split into five cross-validation folds with an 80:20 train-validation split each. Both training and validation were run with 25 patches of overlap between consecutive context windows. Results in this section are mean patch-by-patch performances for all context windows with standard deviations over the 5 validation folds.

The pretrained models were evaluated for sleep staging using a linear classifier. They were compared to two supervised-only models trained from scratch. The supervised models were of the same architecture as the pretrained models but with half the number of layers, attention heads, and embedding dimensionality. This was done to mitigate overfitting. One supervised model used only the raw accelerometry signal as input, while the other had an accelerometry decomposition layer first. This layer first extracts the respiratory and pulse signals, as well as an overall activity signal, calculated as the absolute value of the cross-axial Euclidian-norm-minus-one, and stacks these with the raw signal for a total 8 channel input consisting of the 3 axes from the accelerometry, and respiratory signals, and one from of the pulse and activity signals.

### Apnea Severity Evaluation

We derived 12 summary statistics from the output probabilities from the respiratory event detector trained on the internal training sets to train the random forest apnea severity classifiers. These included:

- Median, Mean and Variance.
- 10^th^, 25^th^, 75^th^, and 90^th^ percentiles.
- Minimum and Maximum.
- Skew and Kurtosis.
- RE decision threshold crossing rate.

The same summary statistics were derived from the sleep stage classification head along with:

- Sleep stage transition probabilities.
- Percentage of time spent in each stage.
- Total sleep time.
- Stage-wise respiratory event rates.
- Stage-wise mean respiratory event probabilities.
- Correlations between stage– and respiratory event probabilities.
- Mean RE probability in four epochs surrounding stage onsets and offsets. This resulted in a total of 117 features.

## Data Availability

Access to the UK Biobank data can be obtained by application.
The data for the Newcastle cohort can be downloaded at https://zenodo.org/records/1160410#.Y-O65i-l1qs.
Data from the DREAMT cohort can be downloaded at: https://physionet.org/content/dreamt/2.1.0/
Access to data from the remaining cohorts can be requested by contacting the corresponding institutes.

https://zenodo.org/records/1160410

https://physionet.org/content/dreamt/2.1.0/

## Supplementary

### Physiological signal extraction pipeline validation

Relevant frequency ranges for respiration and pulse signals were determined by examining frequency power spectra in nighttime patches with signal standard deviations < 0.01 (Supplementary Fig. 1).

To validate our PyTorch implementation of pulse and respiration signal extraction, we used the TBI training set. The frequency bin with the maximum amplitude in the extracted pulse and respiration spectrograms were used as estimates of heart rate and respiratory rate, respectively, and the same was done with the electrocardiogram and thoracic respiratory inductance plethysmography signals within the same frequency ranges (Supplementary Fig. 2). The frequency ranges considered for heart rate (0.6 – 1.7 Hz) and respiratory rate (0.1 – 0.6) allow maximum absolute errors of ∼65 and 30 BPM, respectively. We report the fraction of epochs with absolute errors < 5 BPM for respiratory rate and < 10 BPM for heart rate. The thresholds were chosen to give similar expected values of 0.13 and 0.12 for random guessing.

Overall, we found respiratory rate to be more robustly estimated than heart rate across all conditions (Supplementary Tables 1, 2). Applying a standard deviation threshold of < 0.015, which was used for model pretraining, increased accuracy of both heart– and respiratory rate estimation (Supplementary Table 1).

**Supplementary Figure 1.**
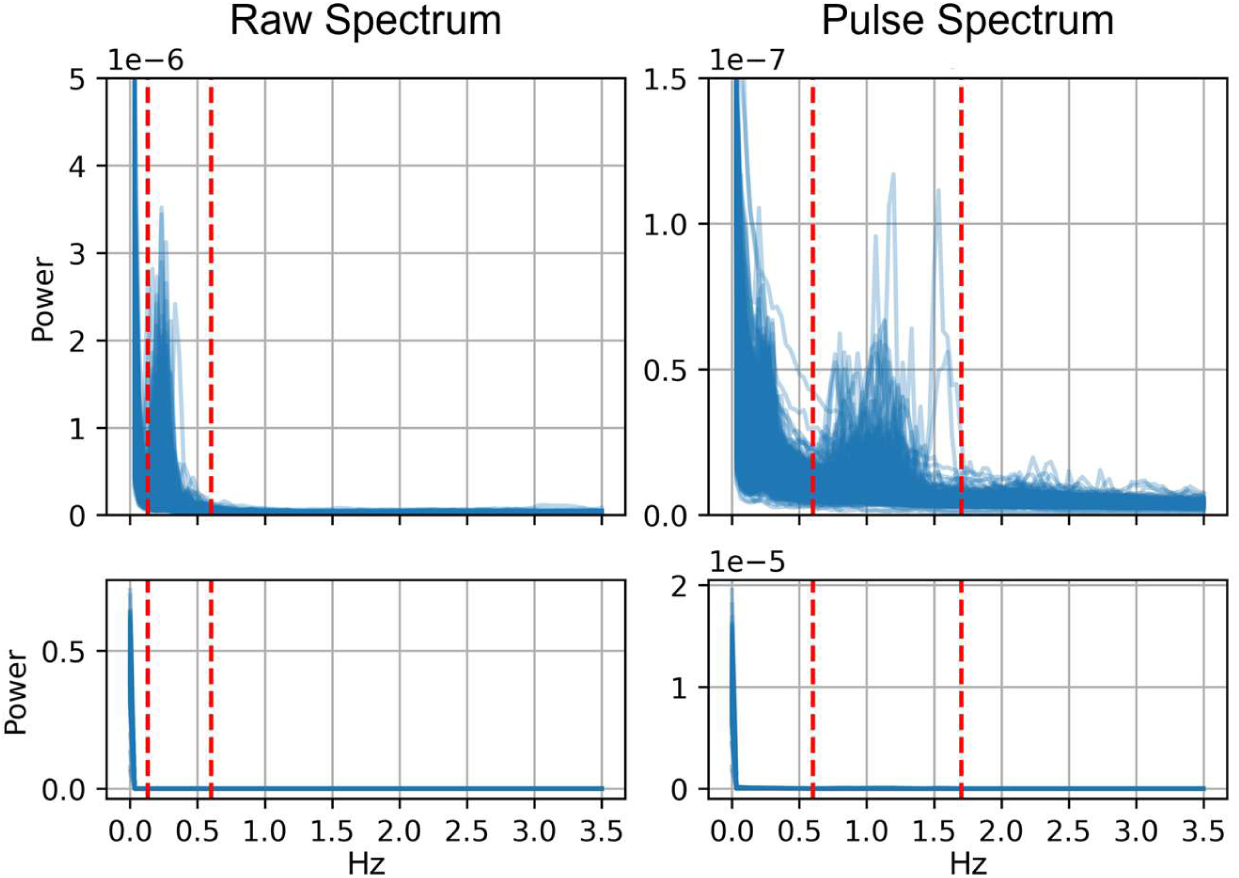
| Average Power Spectra in Quiet Night-Time Patches. Average low-pass (< 3.5 Hz) spectra of 30 s epochs with standard deviation < 0.01 g in 500 12-hours 9 pm – 9 am periods from UK Biobank participants. **Left:** Raw accelerometry signal. Dashed lines show the 0.13 – 0.6 Hz breathing band. **Right:** Derived pulse signal. Dashed lines show the 0.6 – 1.7 Hz pulse-band. **Top:** Zoom-in on the power axis to show the low-amplitude breathing and pulse peaks. **Bottom:** Full power range.

**Supplementary Table I.** Accuracy of accelerometry-based heart rate (HR) and respiratory rate (RR) Estimates Across Sleep Stages.

|  | All | SD < 0.015 | N3 | N2 | N1 | REM | Wake |
| --- | --- | --- | --- | --- | --- | --- | --- |
| **Epochs** | 161851 | 128773 | 31607 | 51395 | 14750 | 23861 | 38781 |
| **Heart Rate** | 0.56 | 0.65 | 0.66 | 0.61 | 0.52 | 0.62 | 0.37 |
| **Resp. Rate** | 0.64 | 0.78 | 0.82 | 0.75 | 0.62 | 0.77 | 0.53 |

**Supplementary Table II.** Accuracy of accelerometry-based heart rate (HR) and respiratory rate (RR) Estimates Conditioned on patch Apnea Occurrence.

|  | No Apnea | Hypopnea | Obstructive/Central |
| --- | --- | --- | --- |
| **Epochs** | 136530 | 13585 | 11736 |
| **Heart Rate** | 0.56 | 0.55 | 0.55 |
| **Resp. Rate** | 0.71 | 0.67 | 0.63 |

Scores are the fraction of 30 s epochs with heart rate and respiratory rate estimation errors < 10 and 5 beats/ breaths per minute, respectively.

For individual sleep stages we found both heart– and respiratory rate to be most inaccurately predicted during wakefulness, followed by N1. Similar accuracies were found for N2 and REM sleep, with highest accuracies found during N3. We also observed that heart rate estimation, while less accurate, was less affected by the occurrence of apneas in a 30 s patch (Supplementary Table 2). The distribution of errors, for all conditions were centered around zero, except periods with standard deviations > 0.15, with a bias for underestimation for both heart– and respiratory rate (Supplementary Fig. 3). This was expected for this simplistic method, as accelerometry spectrograms tend to have more power in lower frequencies (Supplementary Fig. 1).

**Supplementary Figure 2.**
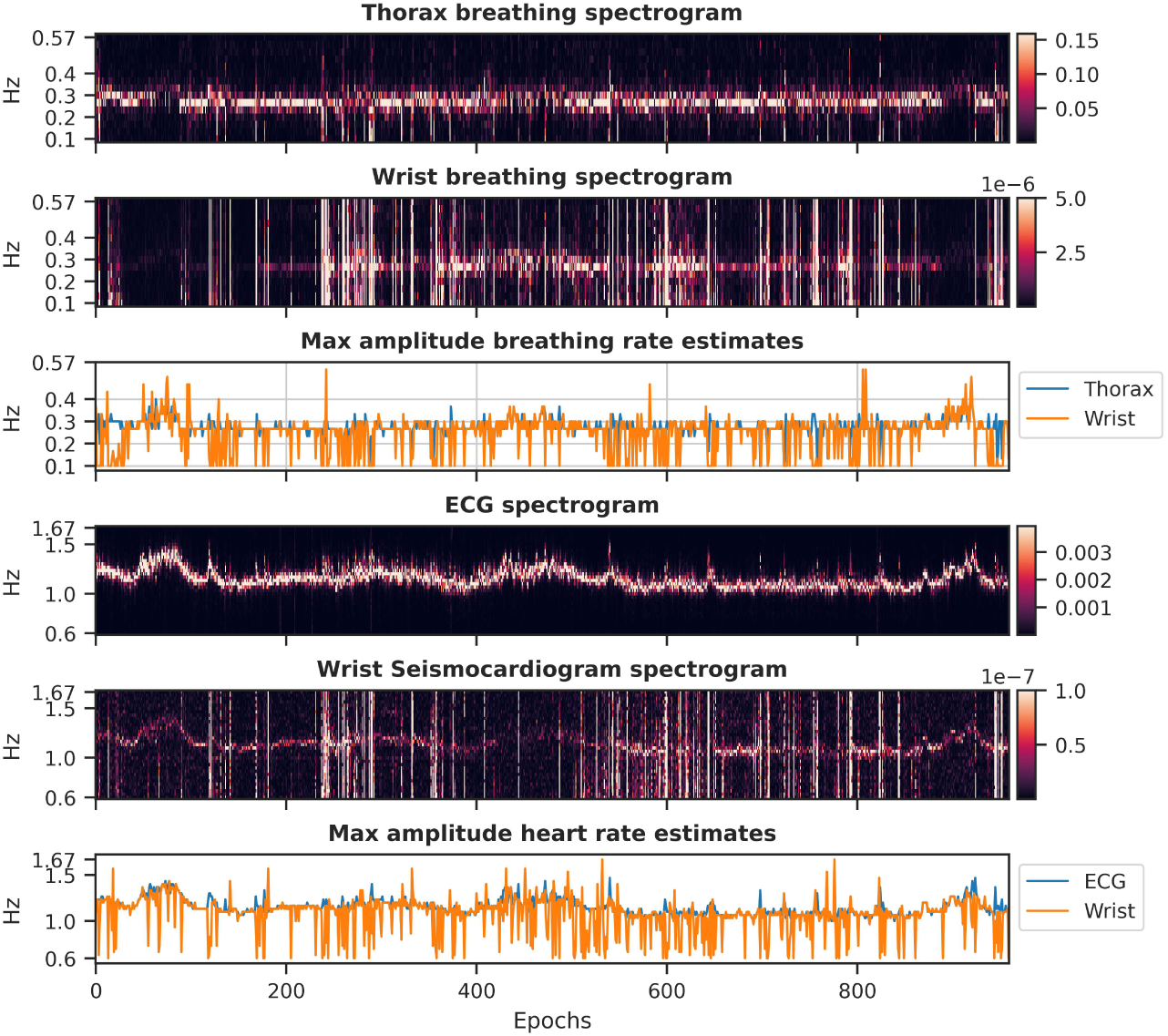
| Estimation of breathing and heart rates from wrist accelerometers. Spectrograms show thoracic respiratory inductance plethysmography and respiratory wrist motion (top) and electrocardiogram and wrist seismocardiogram (bottom) line plots show maximum amplitudes in the two corresponding spectrograms.

**Supplementary Figure 3.**
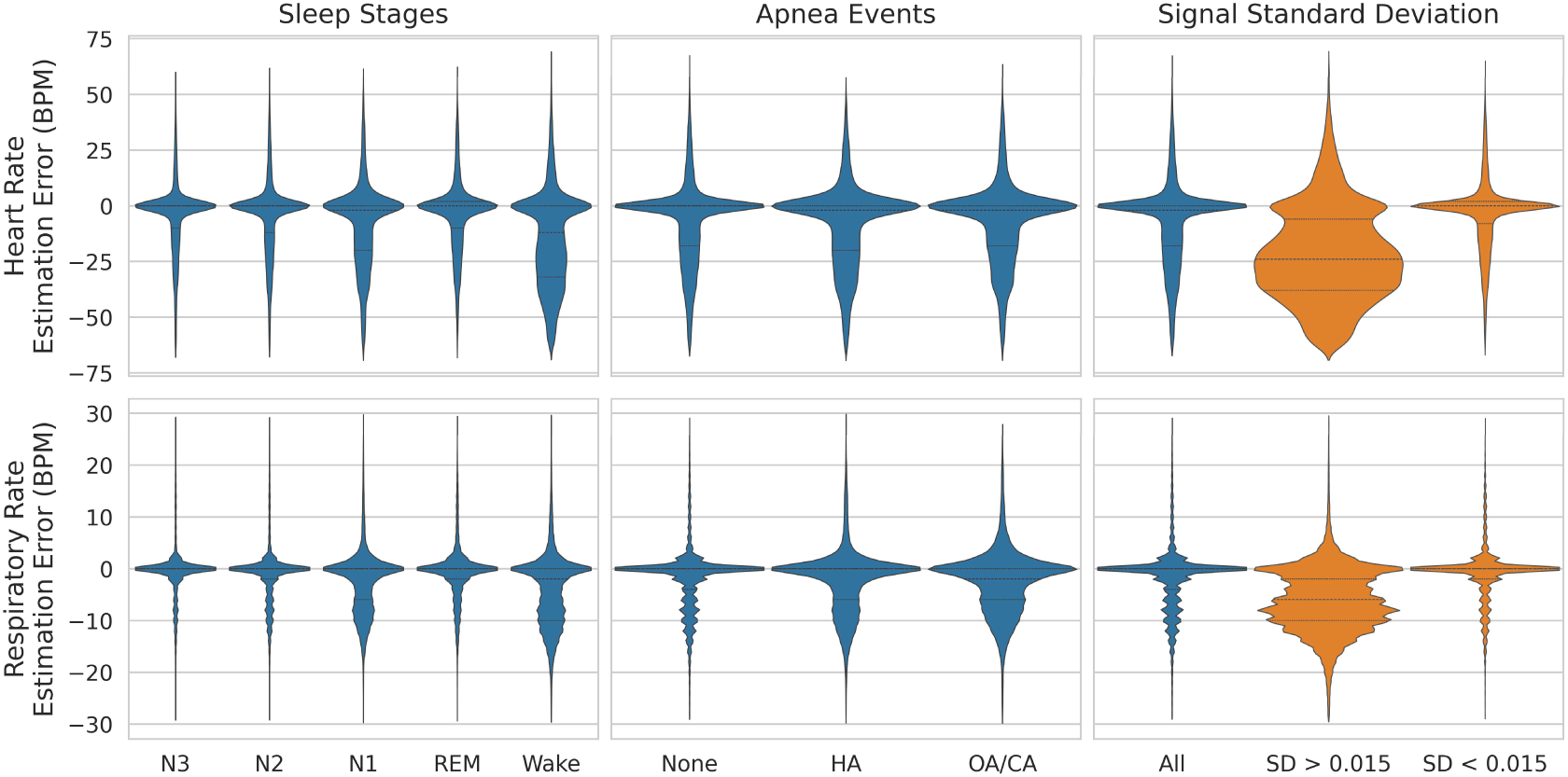
| Distribution of heart– and respiratory rate estimation errors. Errors are from spectrogram max-amplitude estimation across epochs conditioned on sleep stage (left), apnea occurrence (middle), and standard deviation (right).

Consequently, when there is no clear peak, maximum estimations tend toward lower frequencies.

### Classification Head Comparisons

We evaluated four different classifiers trained on the frozen AcceleRest embeddings. The first two were non-sequence models: A single linear layer and a multi-layer perceptron (MLP) with one hidden layer. The other two were sequence models with two bi-directional long short-term memory (LSTM) layers. One of these included a linear pre-LSTM compression layer (LSTM-C), which projected the embeddings to the number of classes before sequence modeling, while the other (LSTM-F) performed sequence modeling directly on the full 256-dimensional pretrained embeddings. We trained all classifiers for 75 epochs and report results from the epoch with the highest macro-average F1 rather than use early stopping (Supplementary Fig. 4a). This was done to investigate overfitting tendencies.

**Supplementary Figure 4.**
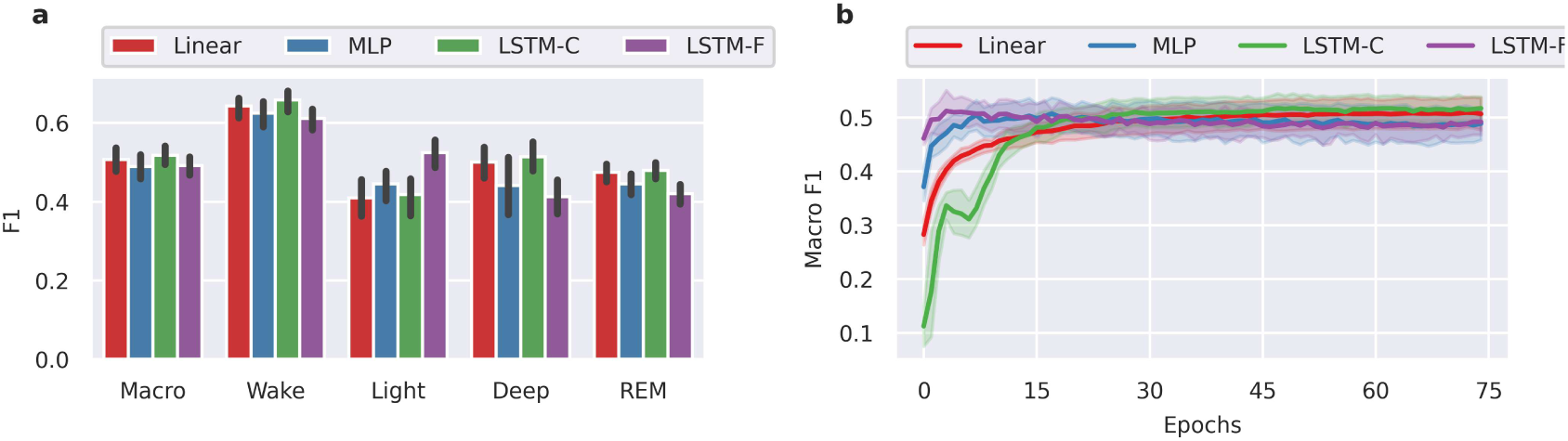
| Classification Head Performance and Learning Curves. **a)** Peak performance of different classification heads trained on AcceleRest embeddings and **b)** validation learning curves. Error bars and learning curve uncertainties are mean ± SD across five folds.

The learning curves (Supplementary Fig. 4b) show that all classifiers except the linear model exhibited overfitting, seen by declining validation performance after a certain epoch. However, LSTM-C did have a slightly higher peak performance compared to the linear classifier, and its performance never dropped below that of the linear classifier. Based on these results linear and LSTM-C classifiers were included for further characterization.

### Data Efficiency and Generalizability

To test the representational power of the AcceleRest embeddings, linear and LSMT-C classifiers were trained on frozen embeddings using 10-100% of the TBI training set, and tested for data efficiency and generalizability on the internal test sets from TBI, DREAMT, STAGES, and Newcastle. The Newcastle cohort was split into two datasets, Newcastle L and R, containing recordings from left and right wrists, respectively. Subject-wise macro F1 scores, on the TBI test set (Supplementary Fig. 5a) show that a linear classifier performed better than LSTM-C when using only 10% (19 individuals) of the training set, with a smaller shift in performance between TBI and the unseen cohorts (Supplementary Fig. 5b). The LSTM-C classifier scaled better on the TBI test set with increasing amounts of training data, surpassing the linear classifier when using 100% (191 individuals) of the TBI training data (Supplementary Fig. 5a).

**Supplementary Figure 5.**
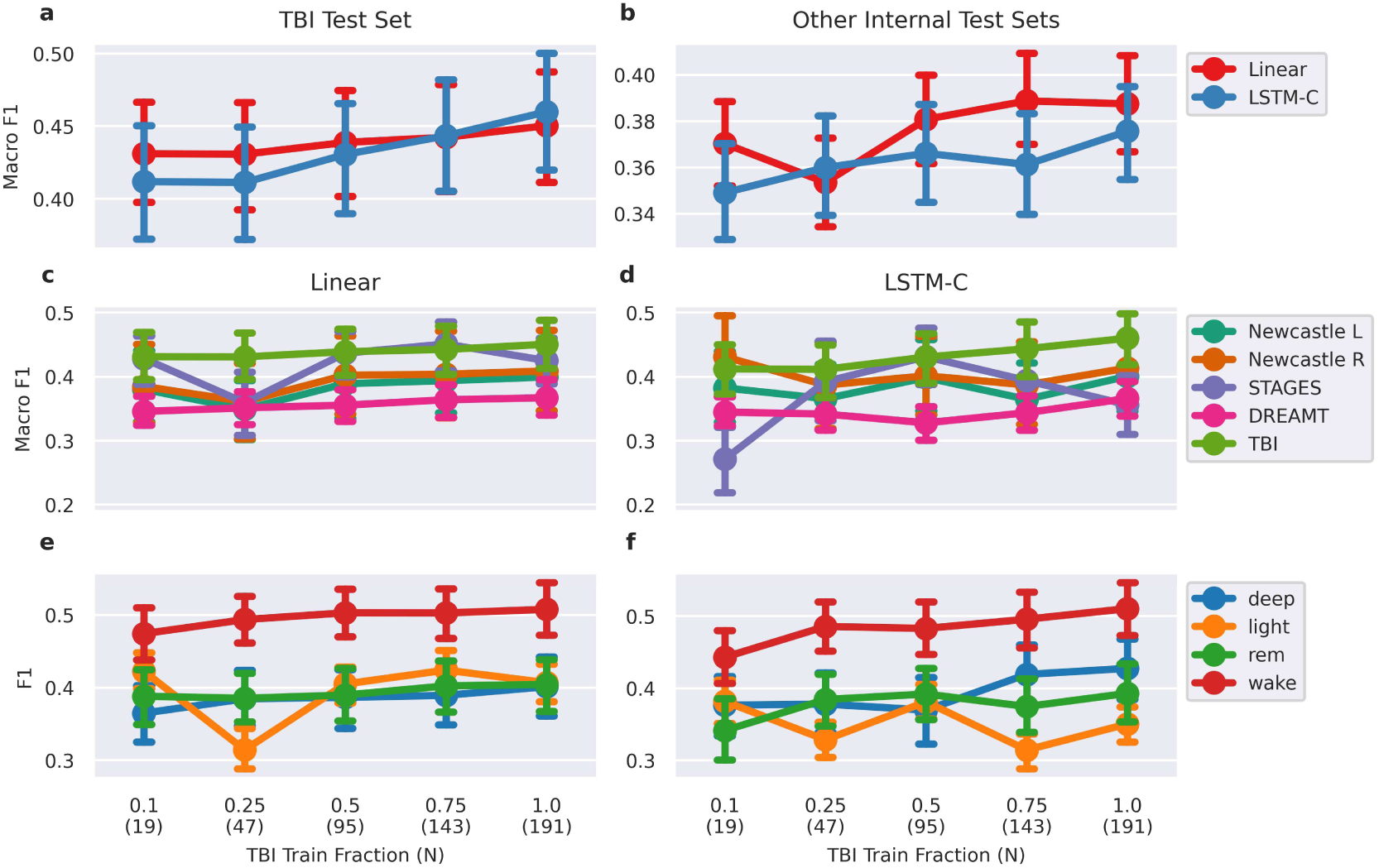
| Cross-cohort performance scaling with the amount of single-cohort training data used for linear and LSTM-C classifiers. Performance is measured as subject-wise macro F1 or F1. Plots show mean performance ± 95% CI. **a)** Performance on the TBI test set for linear and LSTM-C classifiers. **b)** Combined performance on internal test sets from unseen cohorts for linear and LSTM-C classifiers. **c, d)** Linear and LSTM-C classifier performance for each cohort, respectively. **e, f)** Linear and LSTM-C classifer performance for each sleep stage.

However, while performance of both classifiers on the test sets from the unseen internal cohorts scaled with the amount of TBI training data (Supplementary Fig. 5b), the linear classifier performed best for most amounts of training data. Although the difference between the two classifiers when using all the TBI training data was not significant (p = 0.058, subject-wise paired t-test), the linear classifier showed more stable scaling across cohorts (Supplementary Fig. 5c,d) and sleep stages (Supplementary Fig. 5e,f).

To test the effect of including fine-tuning data from target cohorts, we compared performance between linear and LSTM-C classifiers fine-tuned on the frozen AcceleRest embeddings using only the TBI training set or all internal training sets. Larger per-cohort differences were observed for the LSTM-C classifier (Supplementary Fig. 6a,c), suggesting that it learns cohort-wise idiosyncrasies more efficiently than the Linear model. Both models showed increased performance on the STAGES and DREAMT cohorts. Overall, increases were explained mainly by improvements in performance for light sleep and REM sleep (Supplementary Fig. 6d). Looking at the confusion matrices for STAGES when models were trained with only data from TBI (Supplementary 7), we observed a strong bias for overpredicting deep sleep and underpredicting light and REM, especially for the LSTM-C model. This was largely corrected when training on all internal training sets (Supplementary 8).

**Supplementary Figure 6.**
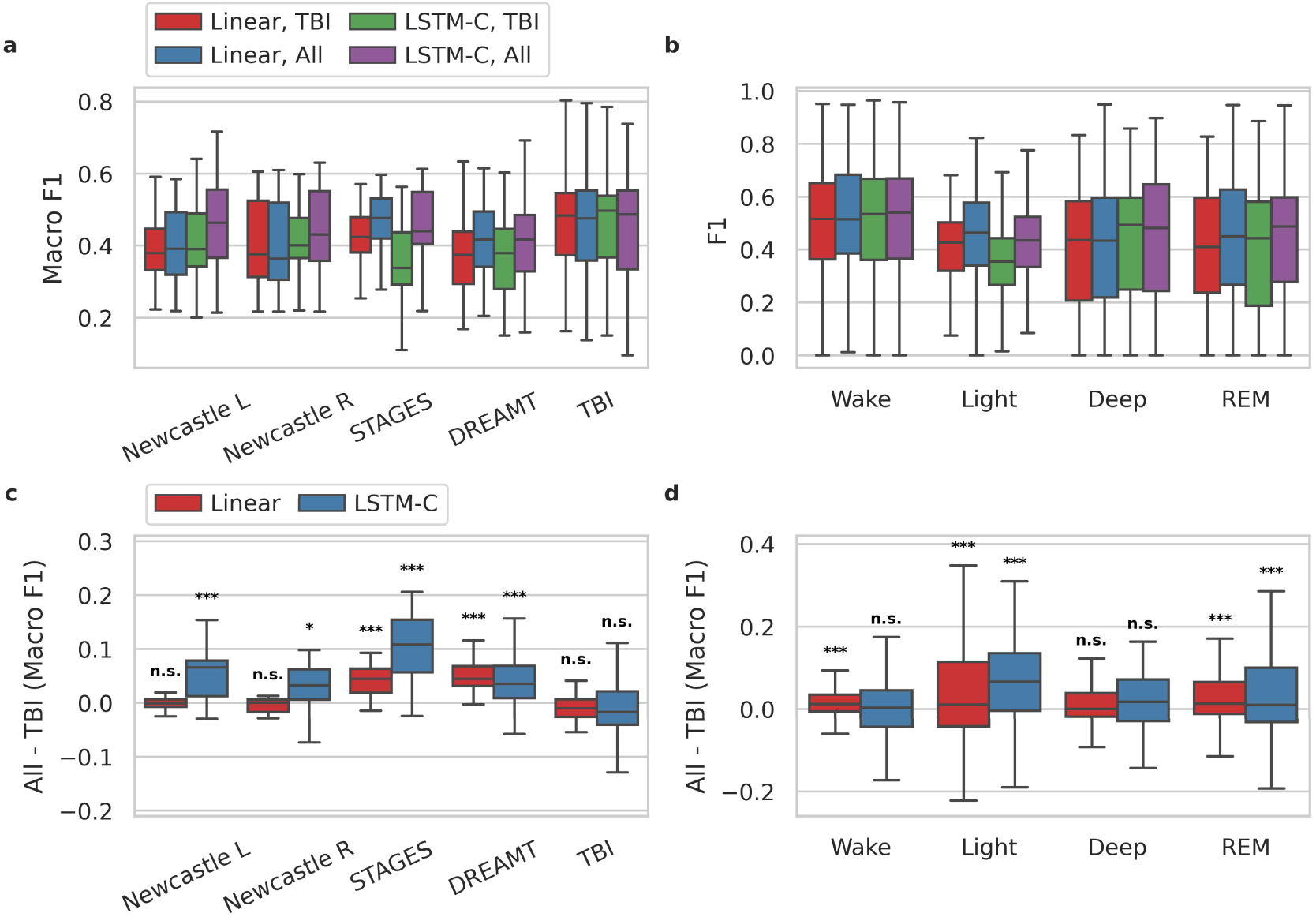
| Effect of Training on Cohort-specific Data. **a)** The subject-wise performance on each cohort and classifier, when finetuned on only the TBI training set or training sets from all internal cohorts. **b)** The subject-wise performance for each sleep stage across cohorts. **c, d)** The subject wise increases in performance when training on all internal training sets vs. TBI only for each classifier. Stars denote p-values from pairwise t-tests.

**Supplementary Figure 7.**
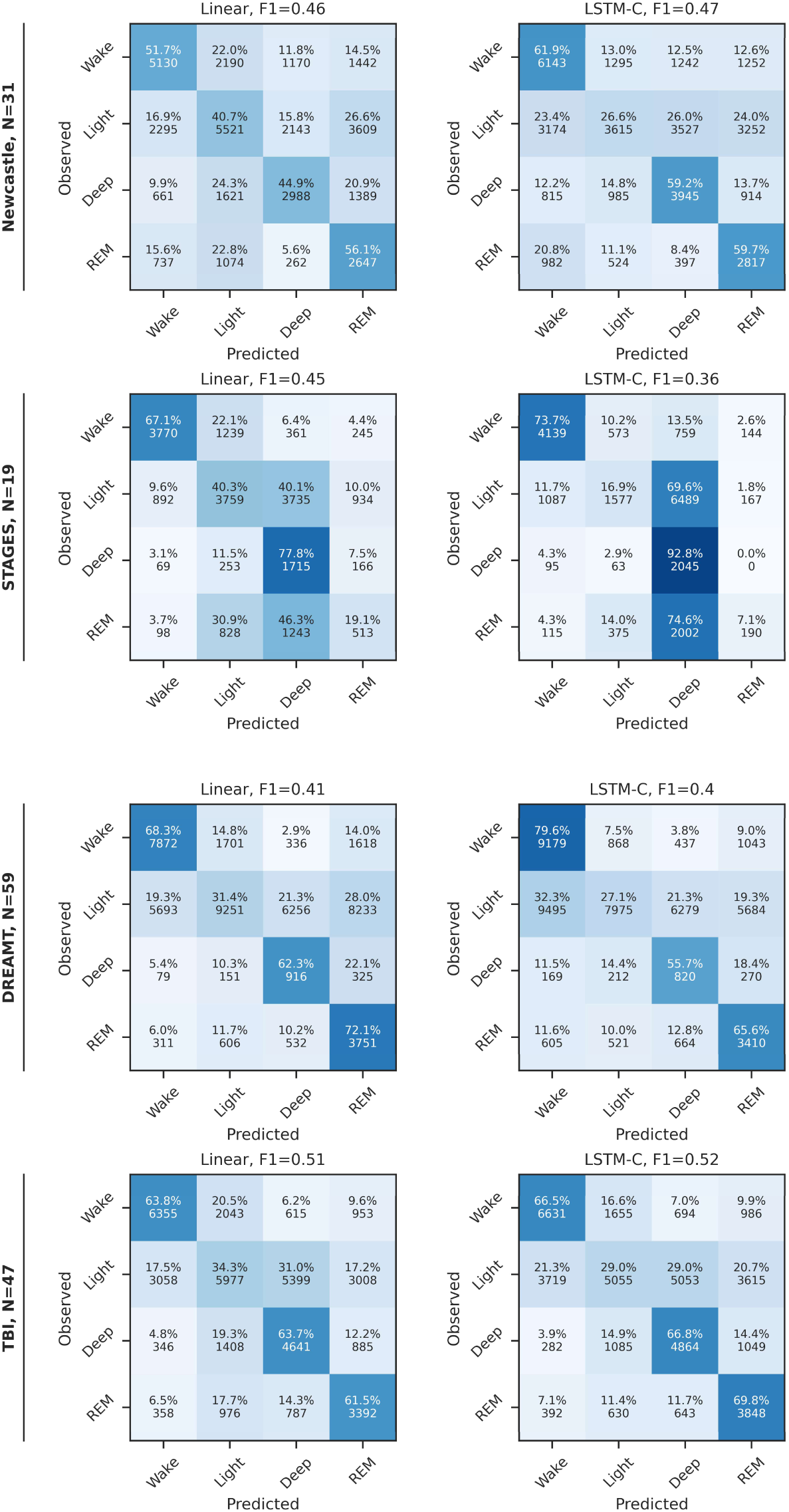
| Confusion matrices for four-class sleep stage classification on internal test sets after training on the TBI training set only. Classification results are for wake, light, deep, and rapid eye movement (REM) classification with a linear (left) or LSTM-C (right) classification heads.

**Supplementary Figure 8.**
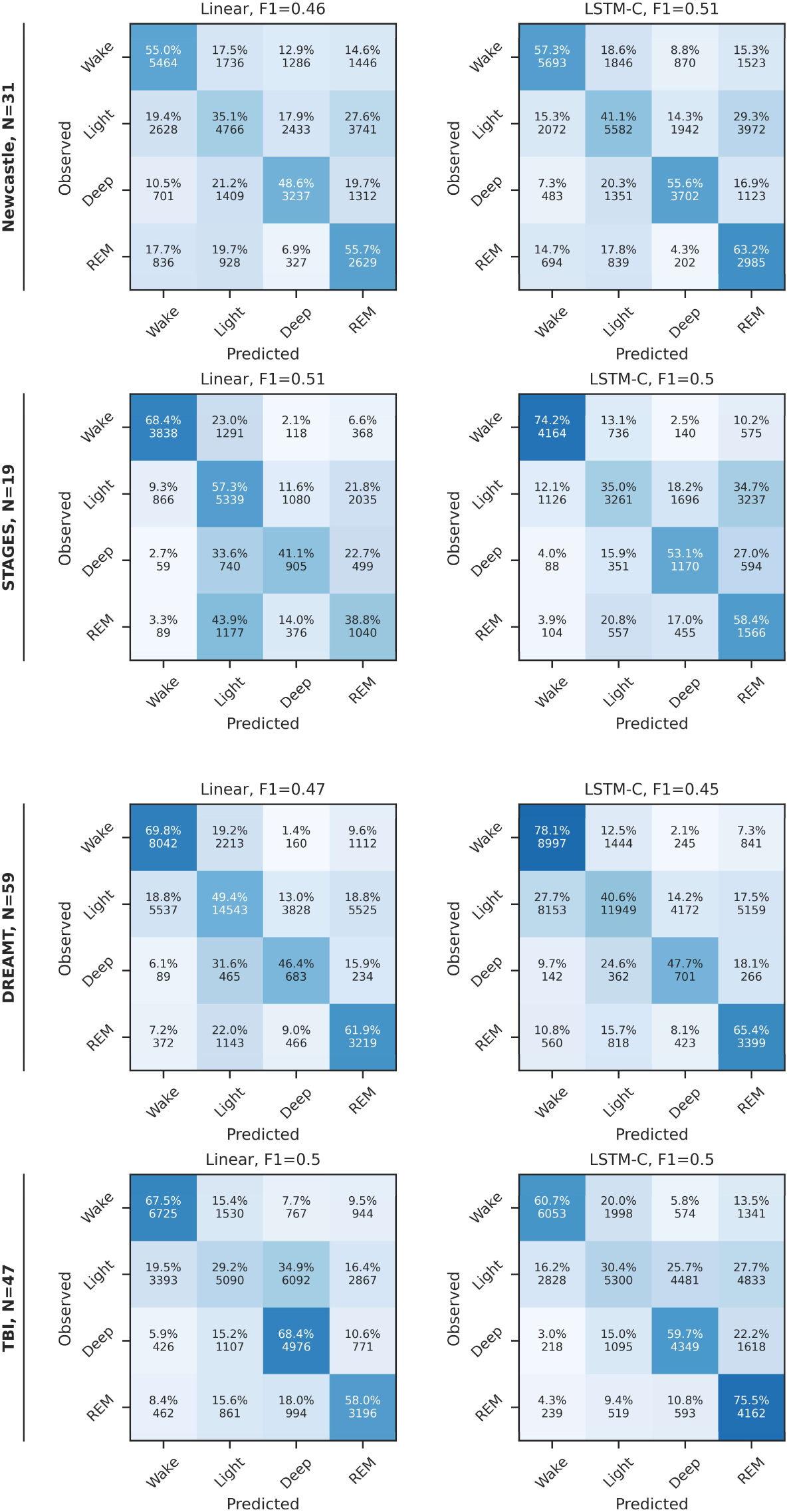
| Confusion matrices for four-class sleep stage classification on internal test sets after training on all internal training sets. Classification results are for wake, light, deep, and rapid eye movement (REM) classification with a linear (left) or LSTM-C (right) classification heads.

### Sleep Summary Metrics

Supplementary tables III and IV show bias and limits of agreement (LoA) from Bland-Altman analysis, as well as the mean absolute error for several sleep summary metrics in the internal and external test sets. For the internal test set evaluation, linear and LSTM-C classifiers were trained on frozen AcceleRest embeddings from all the internal training sets (Supplementary Table 3). For performance on the external test sets, classifiers were trained on all the internal training and test sets (Supplementary Table 4).

**Supplementary Table III.** Internal Test Set Summary Metrics.

| Metric | Classifier | Bias | LoA (lower, upper) | MAE (Mean ± SD) |
| --- | --- | --- | --- | --- |
| TST (Min) | Linear | -7.97 | -161.46, 145.52 | 50.83 ± 60.30 |
| | LSTM-C | -14.66 | -164.71, 135.39 | 55.88 ± 54.52 |
| SE (%) | Linear | -2.57 | -32.85, 27.72 | 10.91 ± 11.27 |
| | LSTM-C | -4.41 | -35.42, 26.61 | 12.32 ± 10.90 |
| REM (Min) | Linear | 41.10 | -76.50, 158.70 | 51.97 ± 51.04 |
| | LSTM-C | 61.15 | -77.39, 199.69 | 67.48 ± 64.88 |
| NREM (Min) | Linear | -49.07 | -193.91, 95.77 | 67.81 ± 57.38 |
| | LSTM-C | -75.81 | -222.05, 70.43 | 90.78 ± 55.62 |
| Deep (Min) | Linear | 32.42 | -109.16, 174.00 | 56.42 ± 55.73 |
| | LSTM-C | 25.97 | -95.82, 147.75 | 49.61 ± 45.69 |
| Light (Min) | Linear | -81.49 | -231.22, 68.24 | 92.52 ± 62.79 |
| | LSTM-C | -101.78 | -251.07, 47.51 | 112.34 ± 59.68 |
| SOL (Min) | Linear | -17.54 | -107.07, 72.00 | 24.77 ± 42.33 |
| | LSTM-C | -12.11 | -95.89, 71.68 | 22.80 ± 38.25 |
| WASO (Min) | Linear | 31.19 | -97.05, 159.43 | 49.78 ± 52.86 |
| | LSTM-C | 27.96 | -90.38, 146.30 | 49.31 ± 44.82 |

LoA: 95% limits of agreement (1.96 SD), MAE: Mean Absolute Error, TST: Total Sleep Time, SE: Sleep Efficiency, REM: Rapid Eye Movement, NREM: Non-REM, SOL: Sleep Onset Latency, WASO: Wakefulness After Sleep Onset.

**Supplementary Table IV.** External Test Set Summary Metrics.

| Metric | Classifier | Bias | LoA (lower, upper) | MAE (Mean ± SD) |
| --- | --- | --- | --- | --- |
| TST (Min) | Linear | 0.80 | -101.60, 103.21 | 33.11 ± 40.79 |
| | LSTM-C | -12.00 | -116.58, 92.58 | 34.09 ± 43.15 |
| SE (%) | Linear | 0.29 | -20.30, 20.89 | 6.54 ± 8.30 |
| | LSTM-C | -2.05 | -22.30, 18.20 | 6.75 ± 8.16 |
| REM (Min) | Linear | 32.46 | -68.71, 133.62 | 47.69 ± 38.33 |
| | LSTM-C | 58.21 | -67.43, 183.84 | 63.63 ± 59.24 |
| NREM (Min) | Linear | -31.65 | -163.08, 99.77 | 53.26 ± 52.06 |
| | LSTM-C | -70.21 | -208.64, 68.23 | 77.81 ± 62.71 |
| Deep (Min) | Linear | 119.76 | -48.38, 287.89 | 131.94 ± 66.12 |
| | LSTM-C | 76.74 | -79.30, 232.78 | 91.42 ± 62.77 |
| Light (Min) | Linear | -151.41 | -295.73, -7.09 | 157.16 ± 60.94 |
| | LSTM-C | -146.95 | -278.48, -15.41 | 149.27 ± 62.33 |
| SOL (Min) | Linear | -11.75 | -75.77, 52.27 | 21.05 ± 27.85 |
| | LSTM-C | 5.08 | -50.48, 60.64 | 16.74 ± 23.64 |
| WASO (Min) | Linear | 15.68 | -58.72, 90.07 | 29.16 ± 29.18 |
| | LSTM-C | 7.78 | -71.21, 86.76 | 25.96 ± 32.08 |

LoA: 95% limits of agreement (1.96 SD), MAE: Mean Absolute Error, TST: Total Sleep Time, SE: Sleep Efficiency, REM: Rapid Eye Movement, NREM: Non-REM, SOL: Sleep Onset Latency, WASO: Wakefulness After Sleep Onset.

**Supplementary 9.**
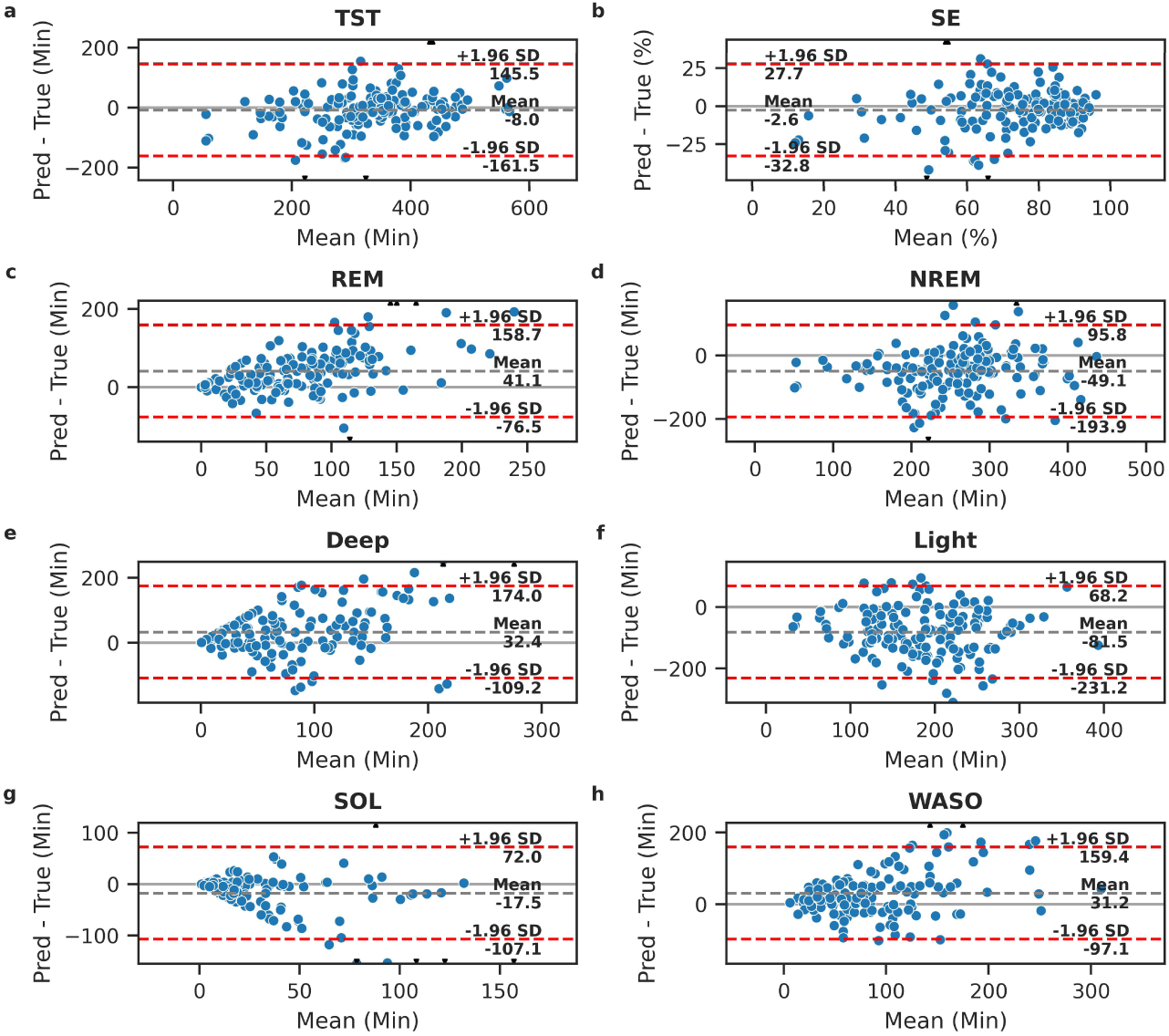
| Sleep Summary Bland-Altman Plots for Linear Classifier on Internal Test Sets. **a)** Total Sleep Time. **b)** Sleep Efficiency. **c)** Total Rapid Eye Movement (REM) sleep. **d)** Total Non-REM sleep. **e)** Total deep sleep. **f)** Total light sleep. g) Sleep Onset Latency. **h)** Wake after sleep onset.

**Supplementary 10.**
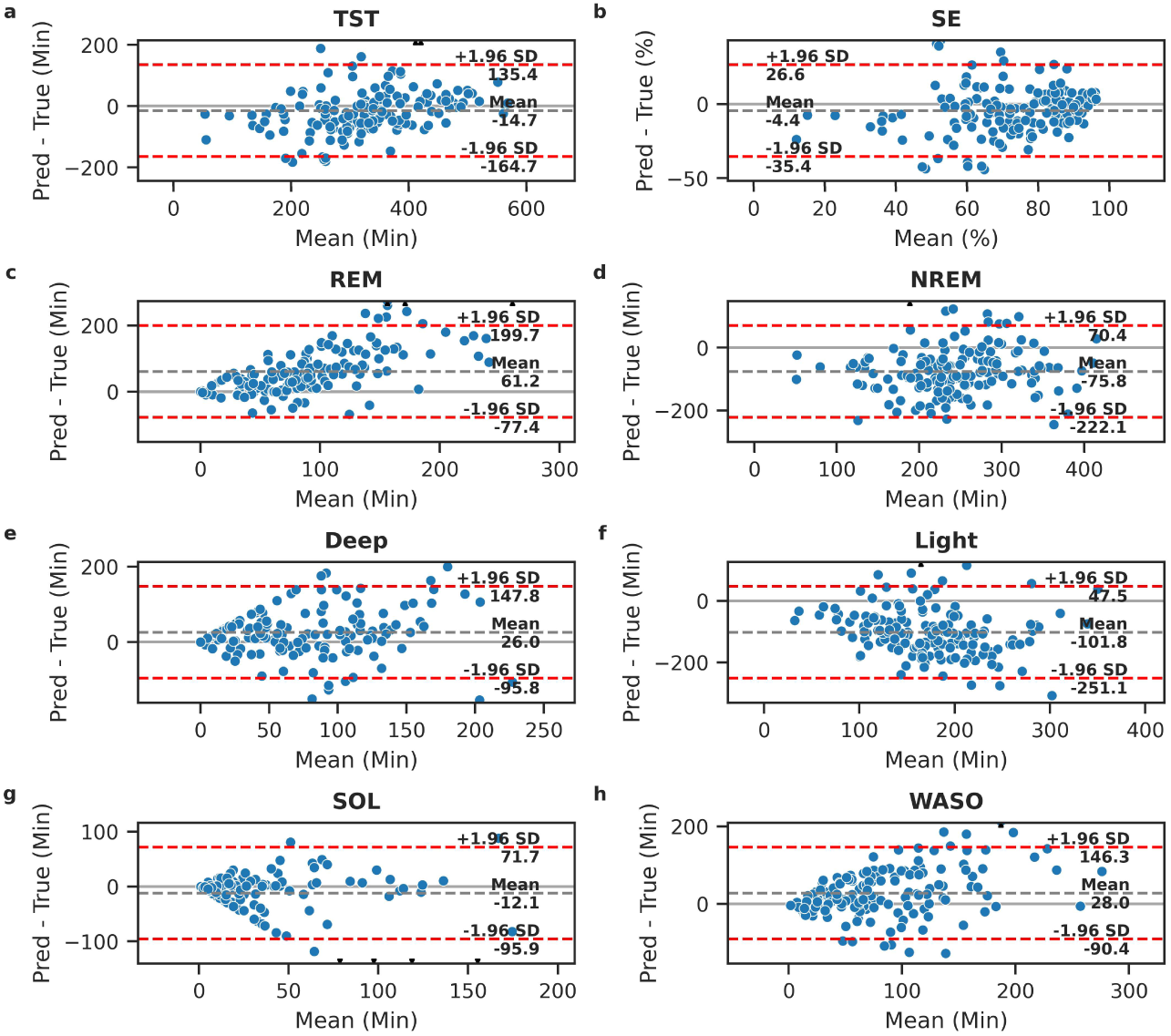
| Sleep Summary Bland-Altman Plots for LSTM-C Classifier on Internal Test Sets. **a)** Total Sleep Time. **b)** Sleep Efficiency. **c)** Total Rapid Eye Movement (REM) sleep. **d)** Total Non-REM sleep. **e)** Total deep sleep. **f)** Total light sleep. g) Sleep Onset Latency. **h)** Wake after sleep onset.

**Supplementary 11.**
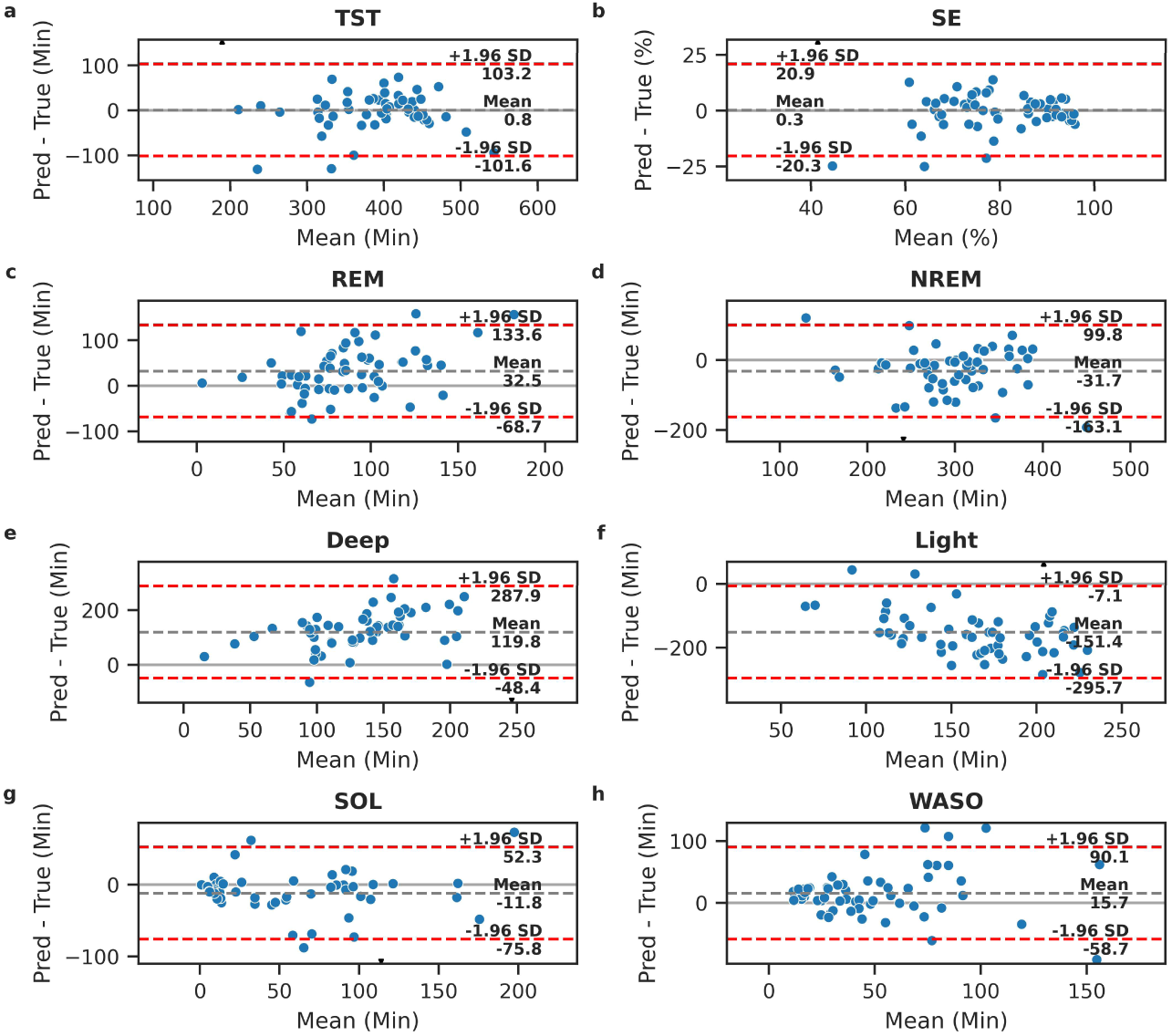
| Sleep Summary Bland-Altman Plots for Linear Classifier on External Test Sets. **a)** Total Sleep Time. **b)** Sleep Efficiency. **c)** Total Rapid Eye Movement (REM) sleep. **d)** Total Non-REM sleep. **e)** Total deep sleep. **f)** Total light sleep. g) Sleep Onset Latency. **h)** Wake after sleep onset.

**Supplementary 12.**
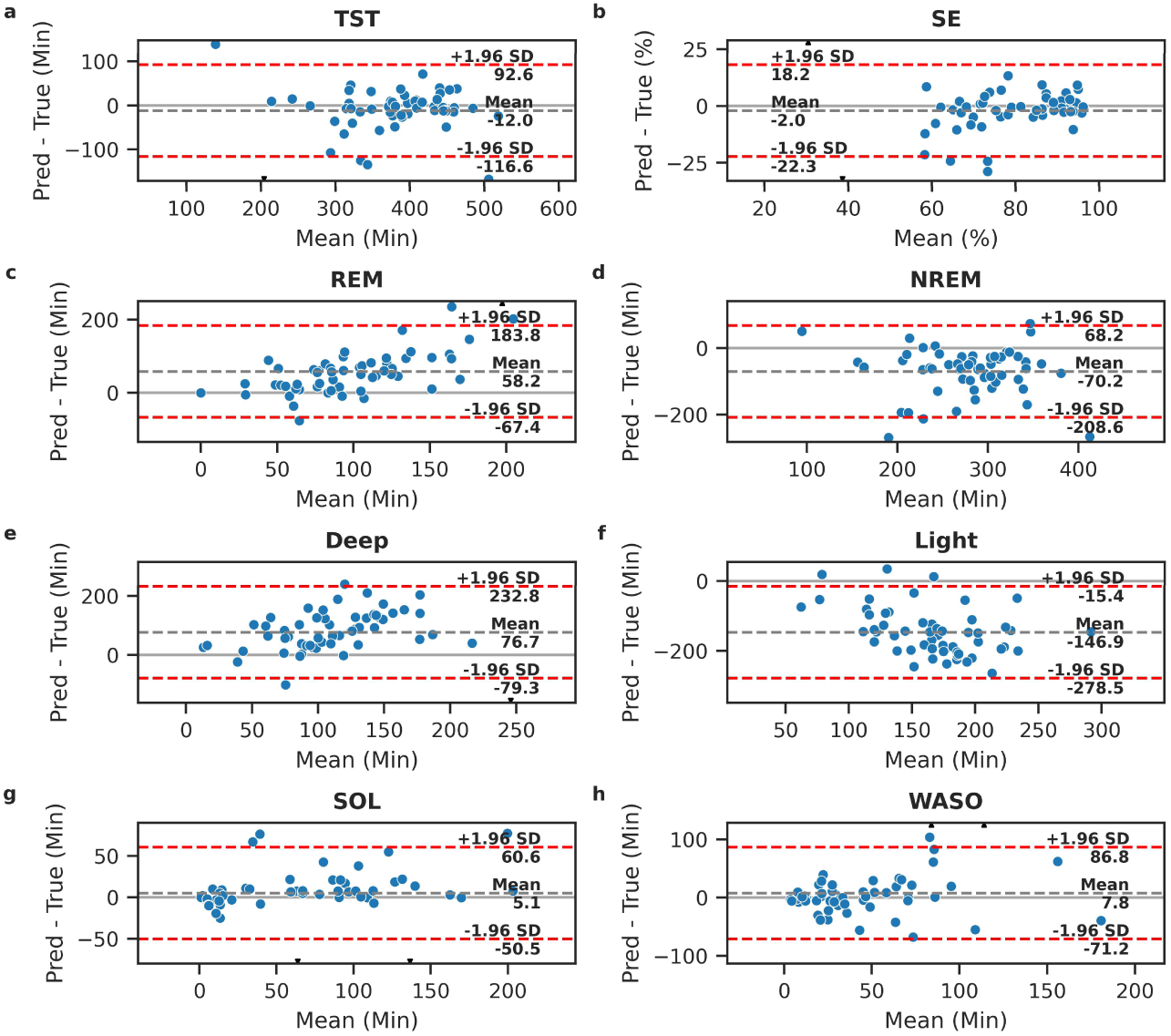
| Sleep Summary Bland-Altman Plots for LSTM-C Classifier on External Test Sets. **a)** Total Sleep Time. **b)** Sleep Efficiency. **c)** Total Rapid Eye Movement (REM) sleep. **d)** Total Non-REM sleep. **e)** Total deep sleep. **f)** Total light sleep. g) Sleep Onset Latency. **h)** Wake after sleep onset.

### Sleep Staging Benchmark vs. SleepNet

First, we compared SleepNet with AcceleRest linear and LSTM-C classifier performance on the external test cohorts, when trained on all internal training and test sets. AcceleRest performance was considerably higher on all reported tasks and metrics for both classifiers compared to SleepNet, with the LSTM-C classifier generally scoring higher than the linear model (Table 5). Comparison based on reported metrics can be prone to aberrant differences due to dataset characteristics. Therefore, we also report a comparison to AcceleRest performance on unseen internal test sets when only training the classifiers on 191 participant nights from the TBI training set (Supplementary Table 1).

**Supplementary Table V.**
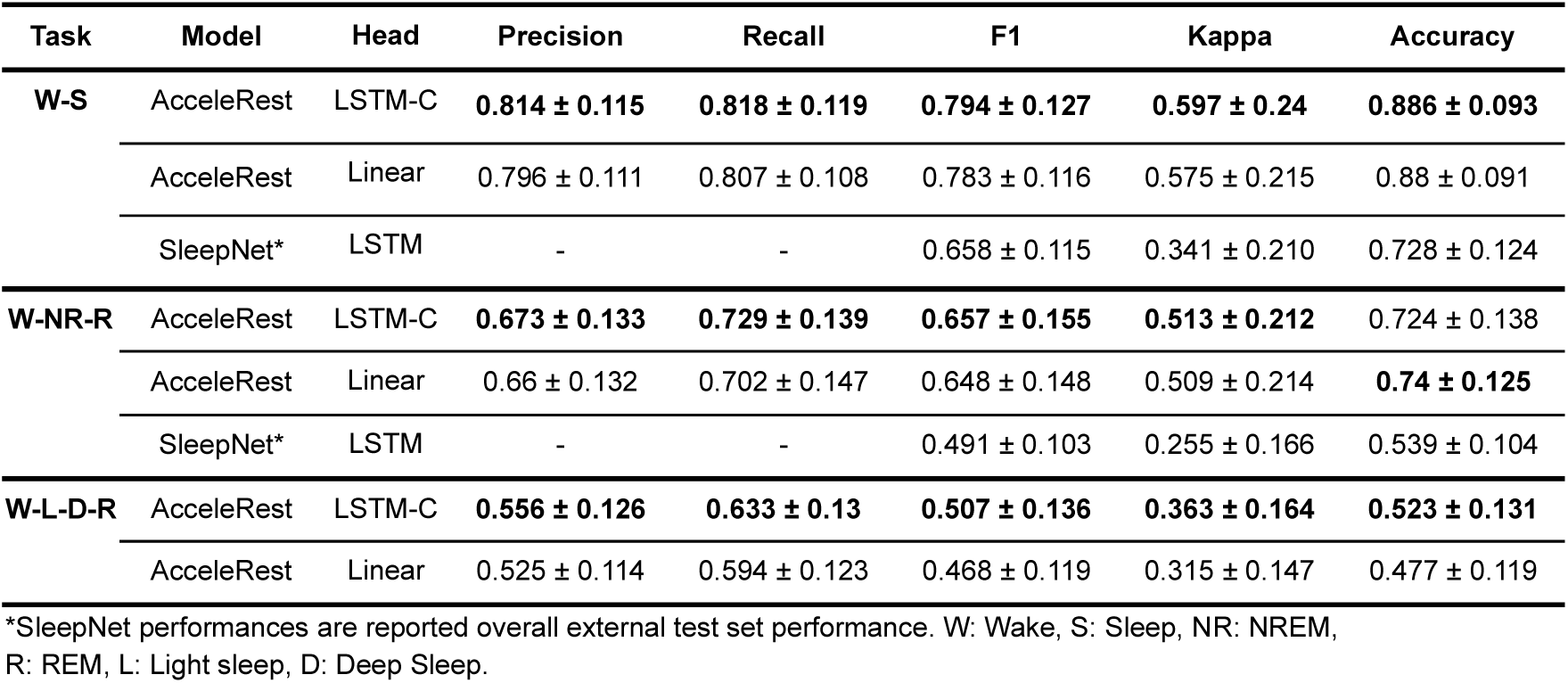
External Test Sets Benchmark.

| Task | Model | Head | Precision | Recall | F1 | Kappa | Accuracy |
| --- | --- | --- | --- | --- | --- | --- | --- |
| **W-S** | AcceleRest | LSTM-C | **0.814 ± 0.115** | **0.818 ± 0.119** | **0.794 ± 0.127** | **0.597 ± 0.24** | **0.886 ± 0.093** |
|  | AcceleRest | Linear | 0.796 ± 0.111 | 0.807 ± 0.108 | 0.783 ± 0.116 | 0.575 ± 0.215 | 0.88 ± 0.091 |
|  | SleepNet* | LSTM | - | - | 0.658 ± 0.115 | 0.341 ± 0.210 | 0.728 ± 0.124 |
| **W-NR-R** | AcceleRest | LSTM-C | **0.673 ± 0.133** | **0.729 ± 0.139** | **0.657 ± 0.155** | **0.513 ± 0.212** | 0.724 ± 0.138 |
|  | AcceleRest | Linear | 0.66 ± 0.132 | 0.702 ± 0.147 | 0.648 ± 0.148 | 0.509 ± 0.214 | **0.74 ± 0.125** |
|  | SleepNet* | LSTM | - | - | 0.491 ± 0.103 | 0.255 ± 0.166 | 0.539 ± 0.104 |
| **W-L-D-R** | AcceleRest | LSTM-C | **0.556 ± 0.126** | **0.633 ± 0.13** | **0.507 ± 0.136** | **0.363 ± 0.164** | **0.523 ± 0.131** |
|  | AcceleRest | Linear | 0.525 ± 0.114 | 0.594 ± 0.123 | 0.468 ± 0.119 | 0.315 ± 0.147 | 0.477 ± 0.119 |

\*SleepNet performances are reported overall external test set performance. W: Wake, S: Sleep, NR: NREM, R: REM, L: Light sleep, D: Deep Sleep.

**Supplementary Table VI.** Held-out Internal Test Set Benchmark with LSTM-C and Linear trained on TBI only.

| Task | Model | Head | Precision | Recall | F1 | Kappa | Accuracy |
| --- | --- | --- | --- | --- | --- | --- | --- |
| **W-S** | AcceleRest | LSTM-C | 0.703 ± 0.117 | **0.733 ± 0.116** | 0.675 ± 0.136 | 0.389 ± 0.226 | 0.763 ± 0.142 |
|  | AcceleRest | Linear | **0.725 ± 0.125** | 0.728 ± 0.122 | **0.698 ± 0.129** | **0.417 ± 0.235** | **0.797 ± 0.13** |
|  | SleepNet* | LSTM | - | - | 0.658 ± 0.115 | 0.341 ± 0.210 | 0.728 ± 0.124 |
| **W-NR-R** | AcceleRest | LSTM-C | **0.579 ± 0.131** | **0.604 ± 0.121** | **0.514 ± 0.128** | **0.334 ± 0.177** | **0.619 ± 0.132** |
|  | AcceleRest | Linear | 0.551 ± 0.115 | 0.589 ± 0.128 | 0.511 ± 0.129 | 0.316 ± 0.178 | 0.61 ± 0.131 |
|  | SleepNet* | LSTM | - | - | 0.491 ± 0.103 | 0.255 ± 0.166 | 0.539 ± 0.104 |
| **W-L-D-R** | AcceleRest | LSTM-C | **0.465 ± 0.119** | **0.52 ± 0.132** | 0.376 ± 0.11 | 0.24 ± 0.126 | 0.451 ± 0.124 |
|  | AcceleRest | Linear | 0.445 ± 0.101 | 0.515 ± 0.131 | **0.387 ± 0.108** | **0.244 ± 0.131** | **0.469 ± 0.115** |

\*SleepNet performances are reported overall external test set performance. W: Wake, S: Sleep, NR: NREM, R: REM, L: Light sleep, D: Deep Sleep.

**Supplementary Table VII.** Internal Test Sets Benchmark with LSTM-C and Linear trained on all internal training sets.

| Task | Model | Head | Precision | Recall | F1 | Kappa | Accuracy |
| --- | --- | --- | --- | --- | --- | --- | --- |
| **W-S** | AcceleRest | LSTM-C | $0.731 \pm 0.12$ | $0.737 \pm 0.117$ | $0.697 \pm 0.123$ | $0.421 \pm 0.218$ | $0.798 \pm 0.119$ |
| | AcceleRest | Linear | $0.737 \pm 0.119$ | $0.738 \pm 0.116$ | $0.707 \pm 0.128$ | $0.437 \pm 0.224$ | **$0.804 \pm 0.127$** |
| | SleepNet* | LSTM | - | - | **$0.749 \pm 0.103$** | **$0.514 \pm 0.186$** | $0.778 \pm 0.096$ |
| **W-NR-R** | AcceleRest | LSTM-C | $0.576 \pm 0.121$ | $0.626 \pm 0.133$ | $0.537 \pm 0.134$ | $0.351 \pm 0.184$ | $0.626 \pm 0.127$ |
| | AcceleRest | Linear | $0.574 \pm 0.116$ | $0.612 \pm 0.13$ | $0.541 \pm 0.132$ | $0.358 \pm 0.189$ | **$0.645 \pm 0.13$** |
| | SleepNet* | LSTM | - | - | **$0.574 \pm 0.111$** | **$0.374 \pm 0.159$** | $0.620 \pm 0.112$ |
| **W-L-D-R** | AcceleRest | LSTM-C | $0.49 \pm 0.115$ | $0.546 \pm 0.136$ | $0.435 \pm 0.13$ | $0.298 \pm 0.155$ | $0.516 \pm 0.126$ |
| | AcceleRest | Linear | $0.474 \pm 0.109$ | $0.531 \pm 0.135$ | $0.427 \pm 0.123$ | $0.293 \pm 0.157$ | **$0.523 \pm 0.129$** |

\*SleepNet performances are reported overall internal test set performance. W: Wake, S: Sleep, NR: NREM, R: REM, L: Light sleep, D: Deep Sleep.

### Sleep Apnea Evaluation

#### Patch-by-patch Respiratory Event Detection

The two-class models had higher recalls for epochs without REs and slightly higher precisions for detecting epochs with REs compared to the three-class models (Supplementary Table 4). While the three-class models had higher recalls for detecting REs, the two-class models had slightly higher F1 scores. The internal test set epoch-by-epoch RE confusion matrices are shown in (Supplementary 19).

**Supplementary Figure 13.**
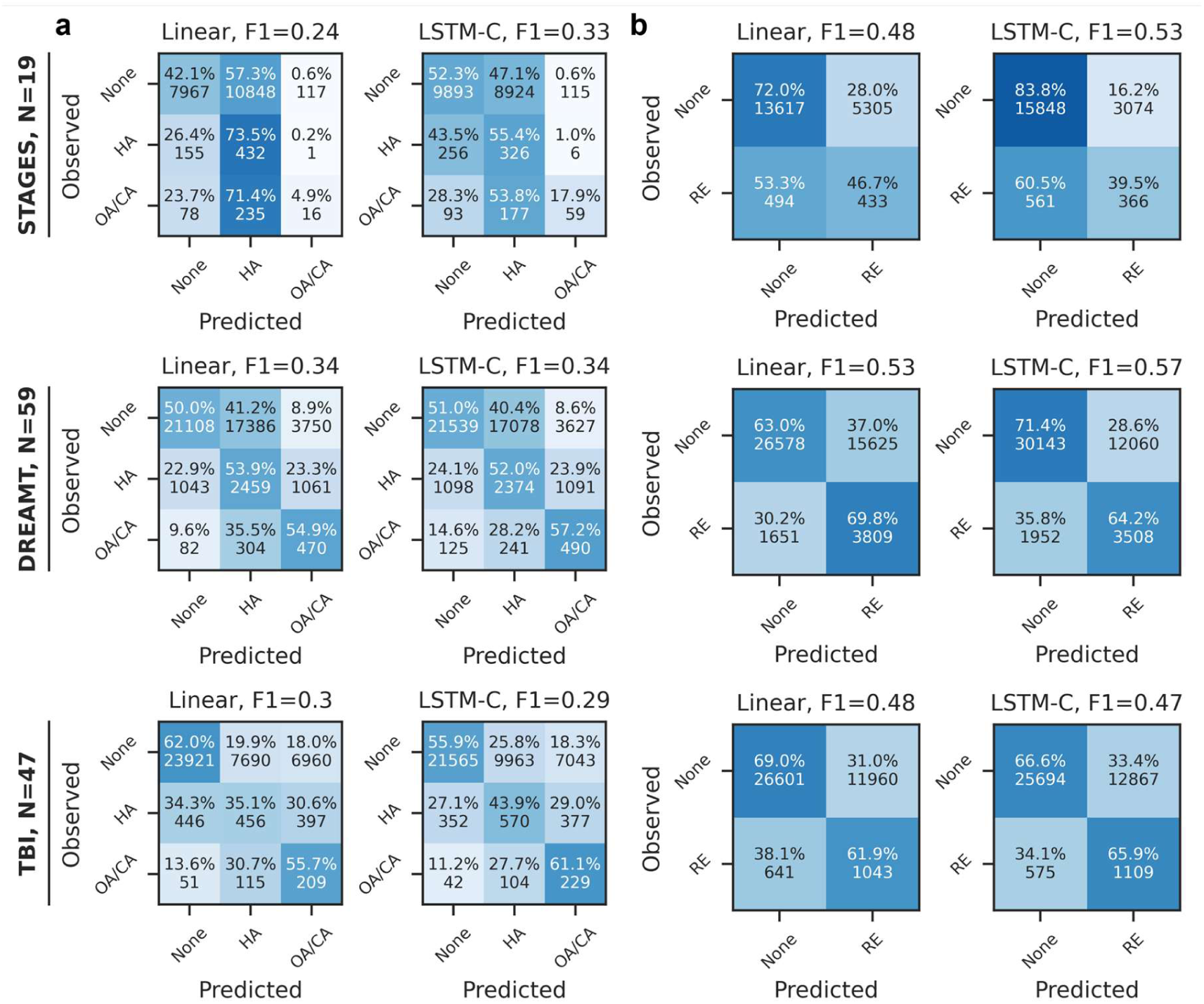
| Three– and Two-Class Epoch-by-Epoch Apnea Event Detection Confusion Matrices. **a)** Three-class model trained for hypopnea (HA) and obstructive/central apnea (OA/CA) classification. **b)** Two-class model trained for combined respiratory event (RE) classification.

**Supplementary Table VIII.** Internal Test Respiratory Event (RE) Detection Performance.

|  |  | Metric | Precision | Recall | F1 |
| --- | --- | --- | --- | --- | --- |
| Class | Training | Classifier |  |  |  |
| None | Three-class | LSTM-C | **0.961 ± 0.077** | 0.521 ± 0.242 | 0.639 ± 0.229 |
|  |  | Linear | 0.96 ± 0.076 | 0.527 ± 0.209 | 0.654 ± 0.196 |
|  | Two-class | LSTM-C | 0.955 ± 0.075 | **0.706 ± 0.246** | **0.781 ± 0.212** |
|  |  | Linear* | 0.953 ± 0.08 | 0.658 ± 0.206 | 0.757 ± 0.182 |
| RE | Three-class | LSTM-C | 0.118 ± 0.125 | 0.721 ± 0.246 | 0.178 ± 0.159 |
|  |  | Linear | 0.12 ± 0.128 | **0.727 ± 0.2** | 0.184 ± 0.163 |
|  | Two-class | LSTM-C | **0.153 ± 0.17** | 0.525 ± 0.309 | 0.189 ± 0.178 |
|  |  | Linear* | 0.132 ± 0.137 | 0.585 ± 0.247 | **0.19 ± 0.165** |

\*Linear two-class model is the model applied for external testing in table 9. Three-class and two-class denotes the grouping used for classifier training.

**Supplementary Table IX.** External Test Set Respiratory Event (RE) Detection Performance.

| Metric | No Event | Respiratory Event |
| --- | --- | --- |
| **AUROC** | 0.719 ± 0.127 | 0.719 ± 0.127 |
| **F1** | 0.854 ± 0.103 | 0.185 ± 0.178 |
| **Precision** | 0.965 ± 0.051 | 0.131 ± 0.146 |
| **Recall** | 0.776 ± 0.137 | 0.489 ± 0.265 |

#### SA Severity and AHI Estimation

We found the directly predicted RE rates to have modest correlation with actual AHIs (Pearson’s R ∼0.5 – 0.55) (Supplementary 21). So rather than using the predicted RE rates to estimate AHI and SA severity directly, we obtained 12 summary metrics (see methods) derived from the epoch-wise estimated RE probabilities us them to train random forest models. We found that the RE detection outputs of the linear classifiers provided better performances on both the SA severity classification (Supplementary 23) and AHI regression (Supplementary 24) compared to the LSTM-C classifiers. The outputs of the two-class model gave the best performance overall with **F1 = 0.51 ± 0.1** (mean ± SD) for SA classification and **R**^2^ **=0.28 ± 0.24** (mean ± SD) for AHI regression. The regression models suffered from overestimation of AHIs in the “normal” range of 0-5 with almost all subjects in this range being predicted to be in the mild to moderate range of 5-30, while the classification models better separated normal and mild severity subjects. Based on this we decided to focus on SA severity classification using the outputs of the linear two-class RE event-detection model.

**Supplementary Figure 14.**
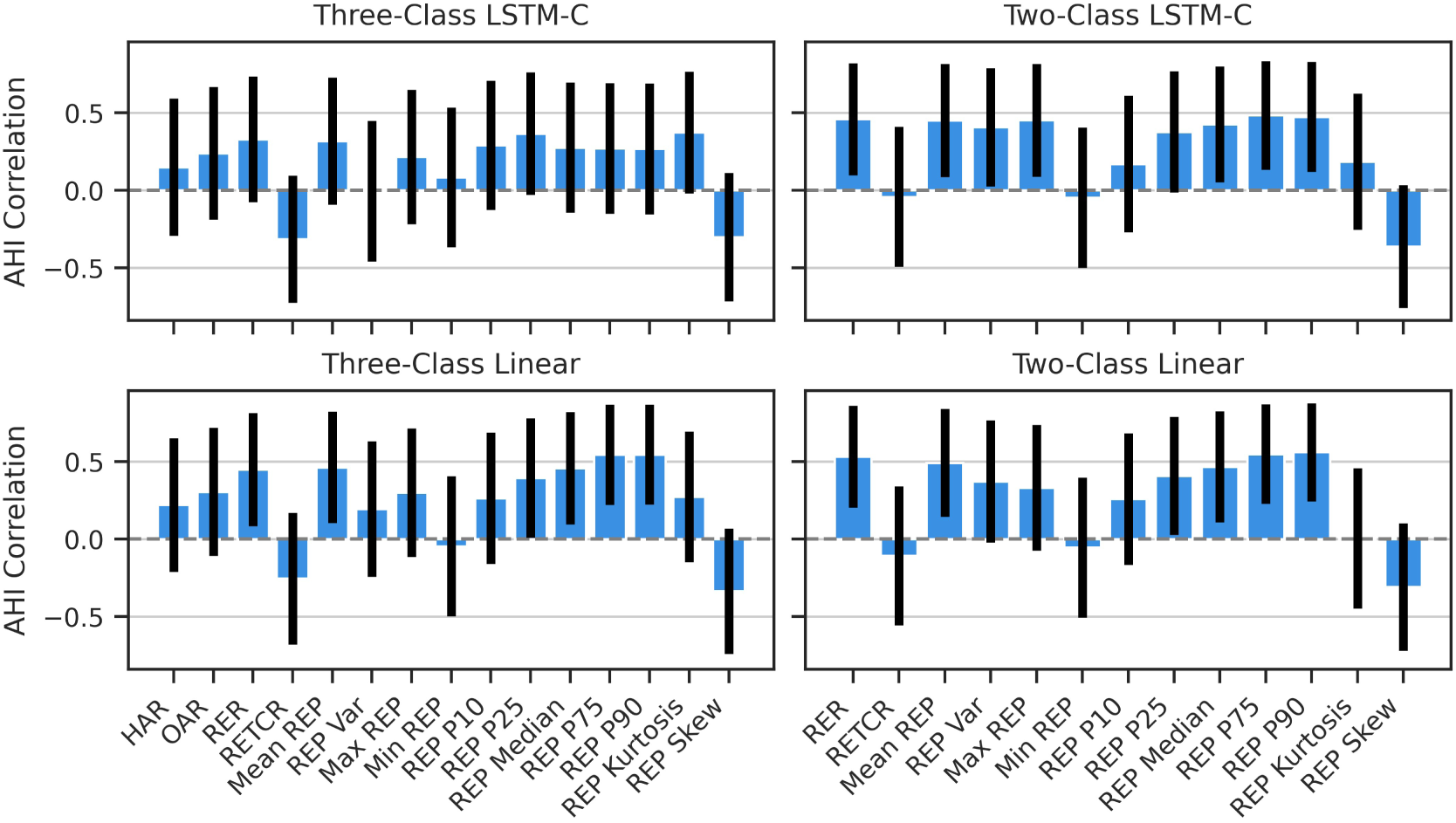
| Correlation of respiratory event detection features and apnea-hypopnea index (AHI). Correlation scores are Pearson’s R. Error bars denote 95% confidence intervals. **HAR**: Hypopnea Rate, **OAR**: Obstructive/Central Apnea Rate, **RER**: Respiratory Event Rate, **RETCR**: Respiratory Event Threshold Crossing Rate, **REP**: Respiratory Event Probability.

**Supplementary Figure 15.**
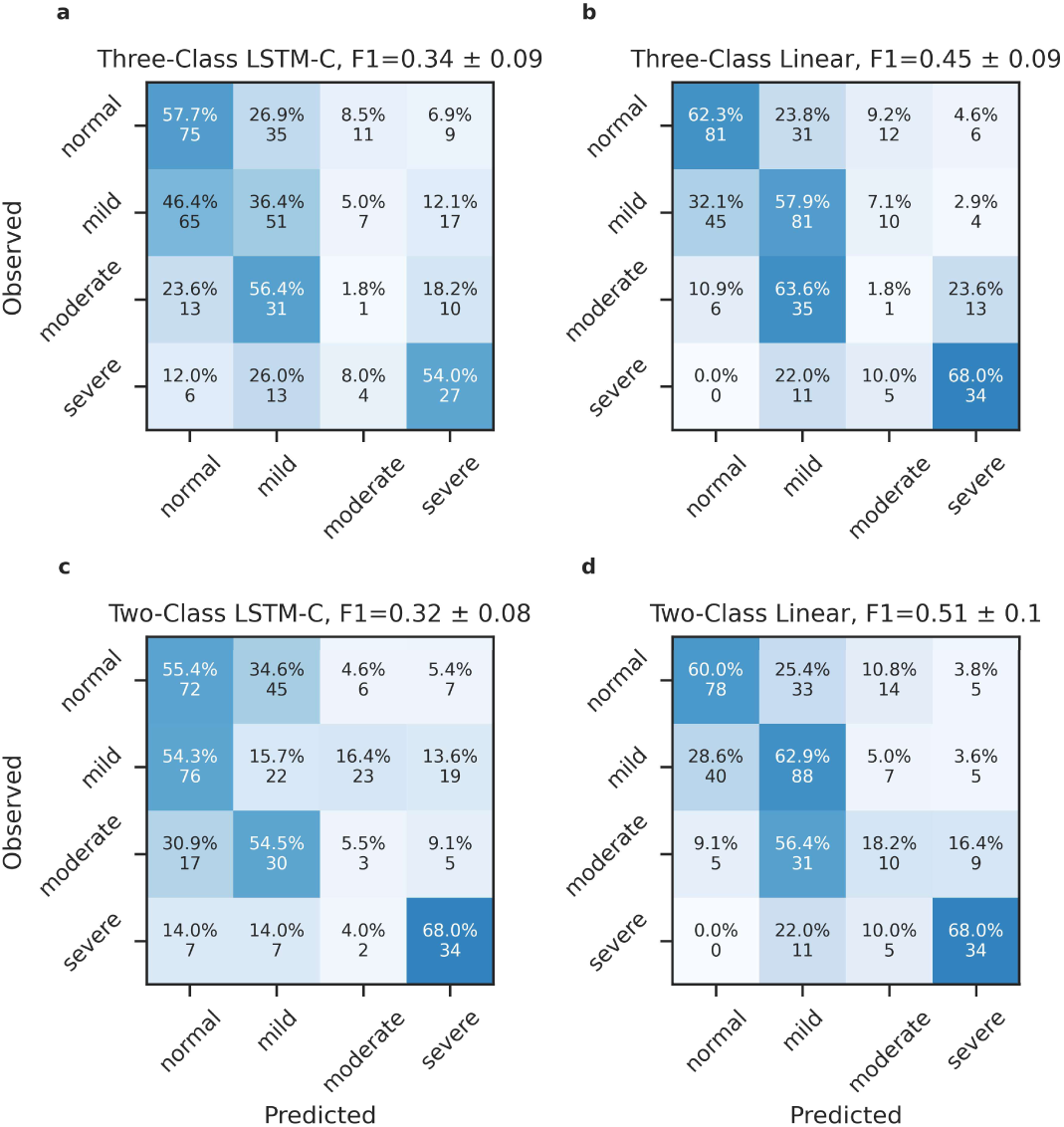
| Sleep apnea severity confusion matrices for for random forest trained on outputs from three-class and two-class respiratory event detection models.

**Supplementary Figure 16.**
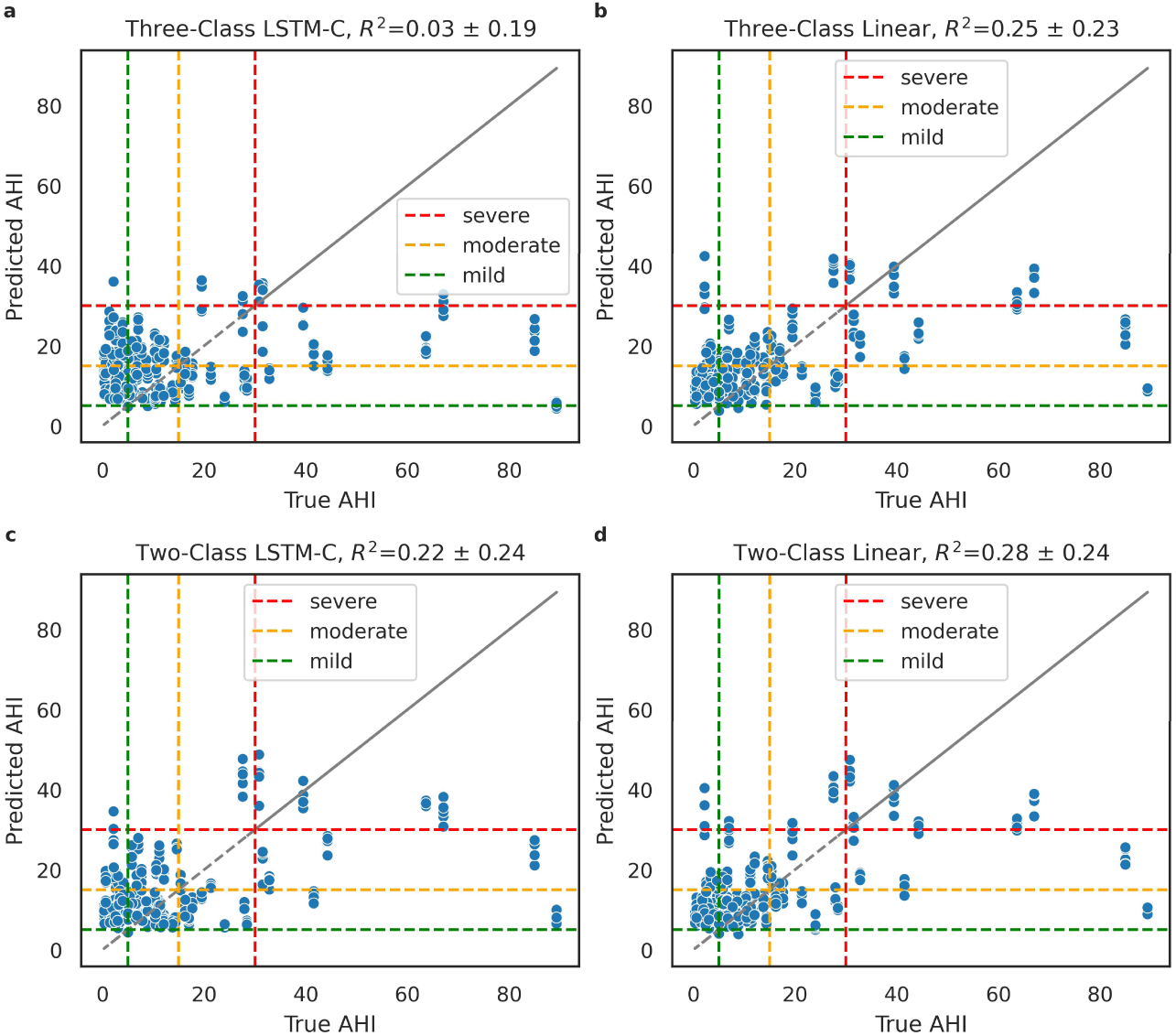
| Apnea-hypopnea index (AHI) random forest regression on outputs from two– and three-class respiratory event detection models. Points in the squares along the diagonal are in the correctly sleep apnea severity range, those above the diagonal have overestimated AHIs, and those below have underestimated AHI.

#### Combined Sleep Staging and Apnea Estimation

To test the effectiveness of combining RE detection and sleep staging for SA classification and to fully utilize the capabilities of AcceleRest. We compared models trained on features from only RE, RE and sleep staging, RE and RE x sleep stage interactions, and the full set of RE, sleep staging, and RE x sleep stage interaction features. We found that inclusion of either the sleep stage features, or RE x sleep stage interaction features alone deteriorated validation performance but including both feature sets boosted performance, with repeated 5-fold CV validation performances of **F1 = 0.59 ± 0.1** (mean ± SD) for the full feature set and **F1 = 0.54 ± 0.12** for only the RE features (Supplementary 25), suggesting that the additional feature sets are only helpful for SA estimation in concert. This could be due to interactions such as the RE rate in a stage with the fraction of time spent in that stage.

**Supplementary 17.**
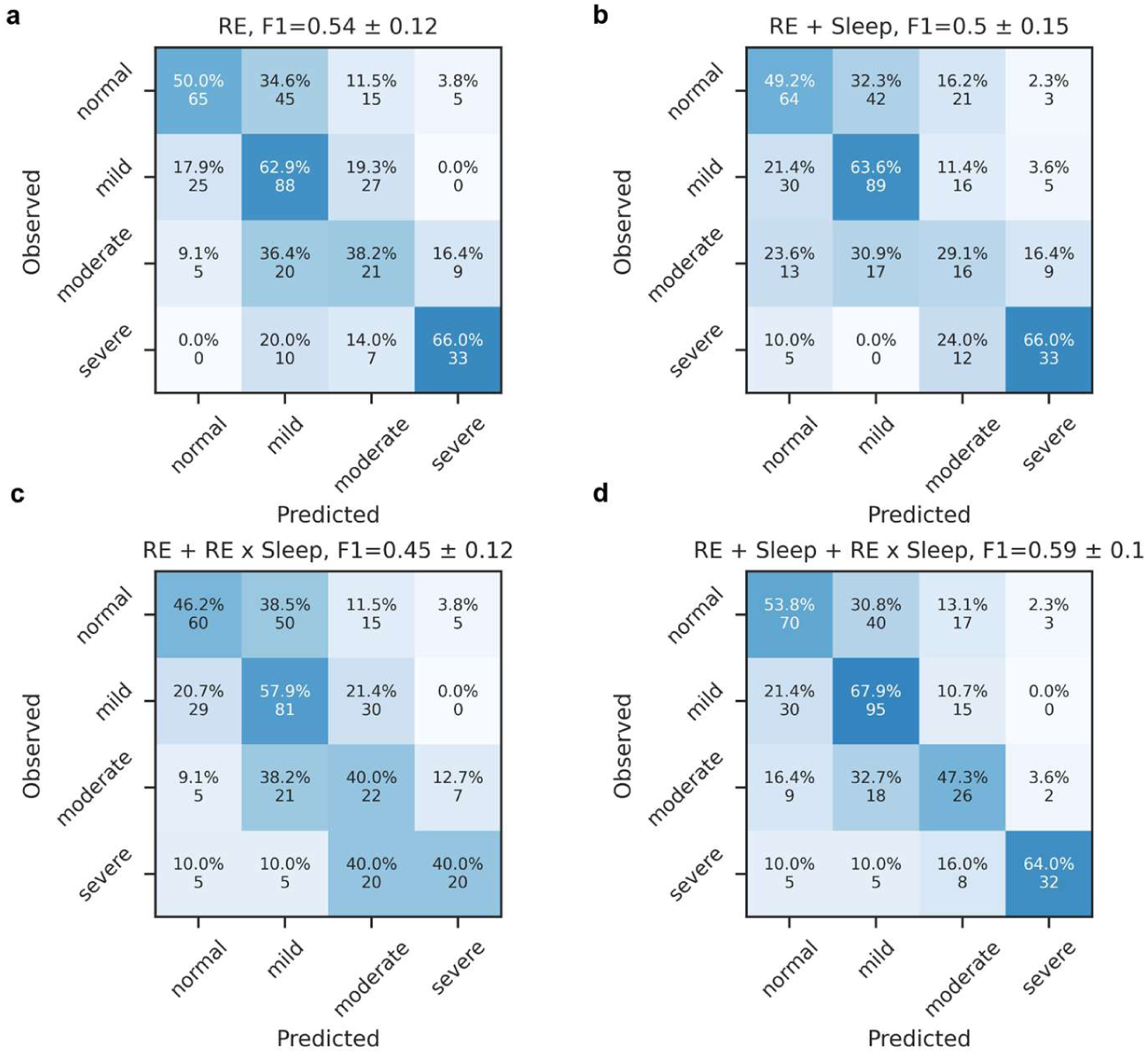
| Sleep apnea severity classification cross-validation confusion. Random forest sleep apnea severity classification with **a)** only respiratory event (RE) feature, **b)** RE and sleep features, **c)** RE and RE-sleep interactions, **d)** RE, sleep and RE-sleep interactions. Numbers reflect 5-fold cross validation repeated 5 times with random splits.

**Supplementary 18.**
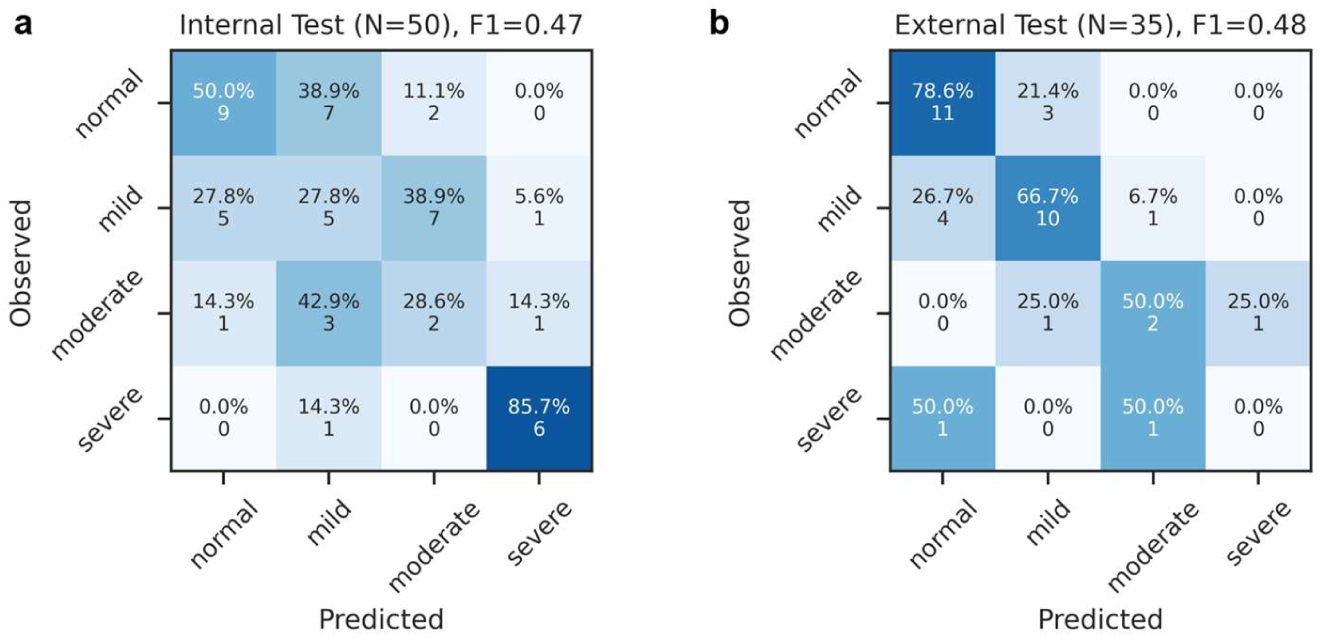
| Internal and external test confusion matrices for random forest sleep apnea severity classifier. Random forest sleep apnea severity classification on **a)** Internal test set, a mix of TBI, STAGES, and DREAMT cohorts and **b)** the Health cohort external test set.

Confusion between the non-severe classes was higher in the internal test set while severe SA was well separated (F1 = 0.47). In the external test set the non-severe classes were comparatively well separated while the two severe SA

**Supplementary 19.**
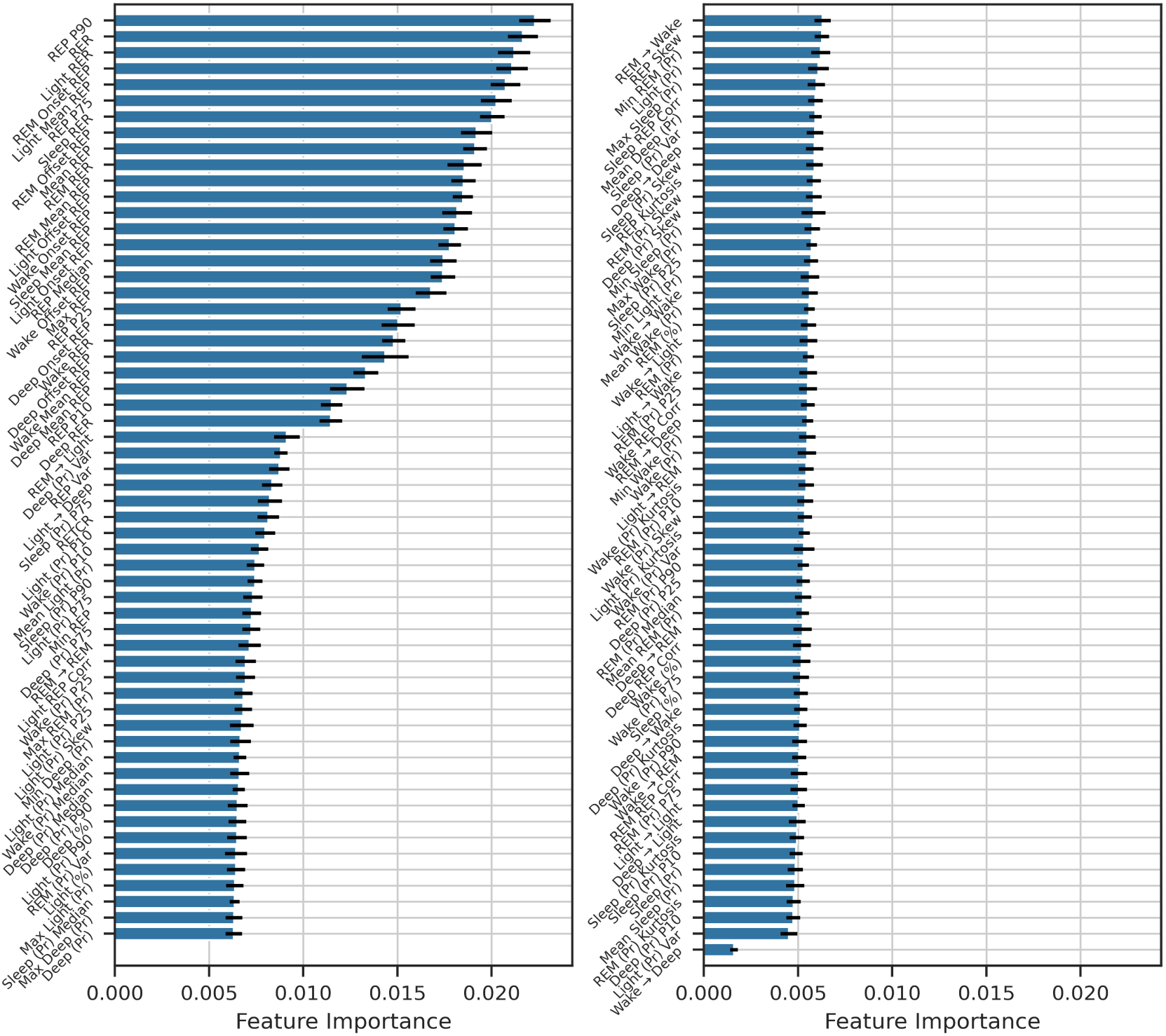
| Respiratory Event Detection Feature Importances for Random Forest SA severity classification. Notable inclusions in the top are overall features such as 90^th^ and 75^th^ percentiles of Respiratory Event (RE) Probabilities (REP P90/P75), RE Rate (RER), combined RE x sleep stage features such as RER during predicted light sleep, RE probability mass around REM offset and onset (REM onset/offset REP), Wake onset REP, Light offset REP.

Looking at the top 15 features by feature importance in the combined RF model with all 117 features (Supplementary 27) we find overall features such as overall RE rate (RER) and RE probability percentiles (REP P90/P75). Notably we also find RE probability mass surrounding stage offset/onsets (stage onset/offset REP) including predicted Light, REM and Wake onsets and offsets. Sleep stage features such as percentages and transition rates generally had lower feature importances. However, since we did not performance an iterative feature exclusion test we cannot tell which features are truly predictive and which are simply correlated with predictive features.

**Supplementary Figure 20.**
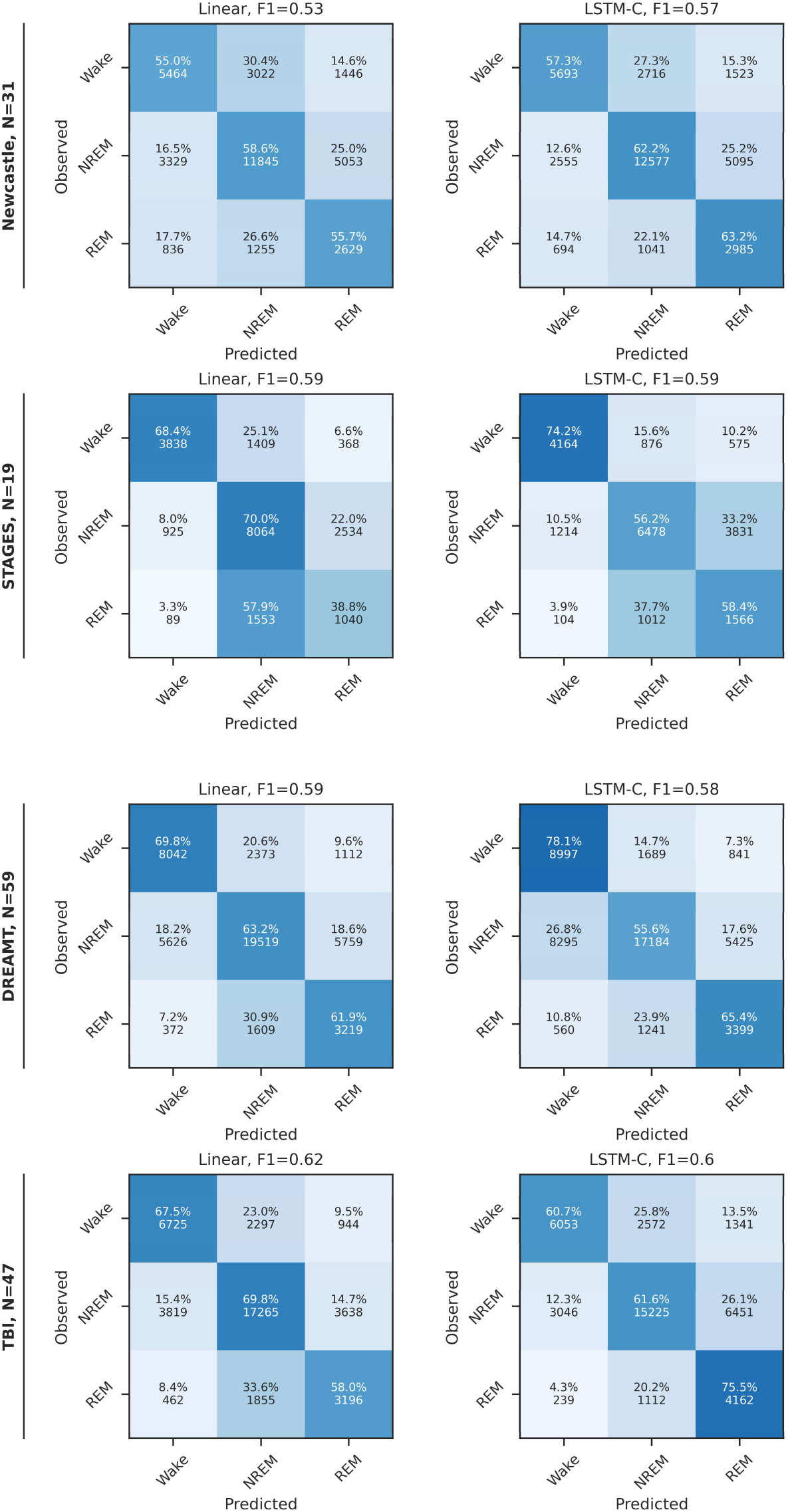
| Wake-NREM-REM confusion matrices for internal test sets.

**Supplementary Figure 21.**
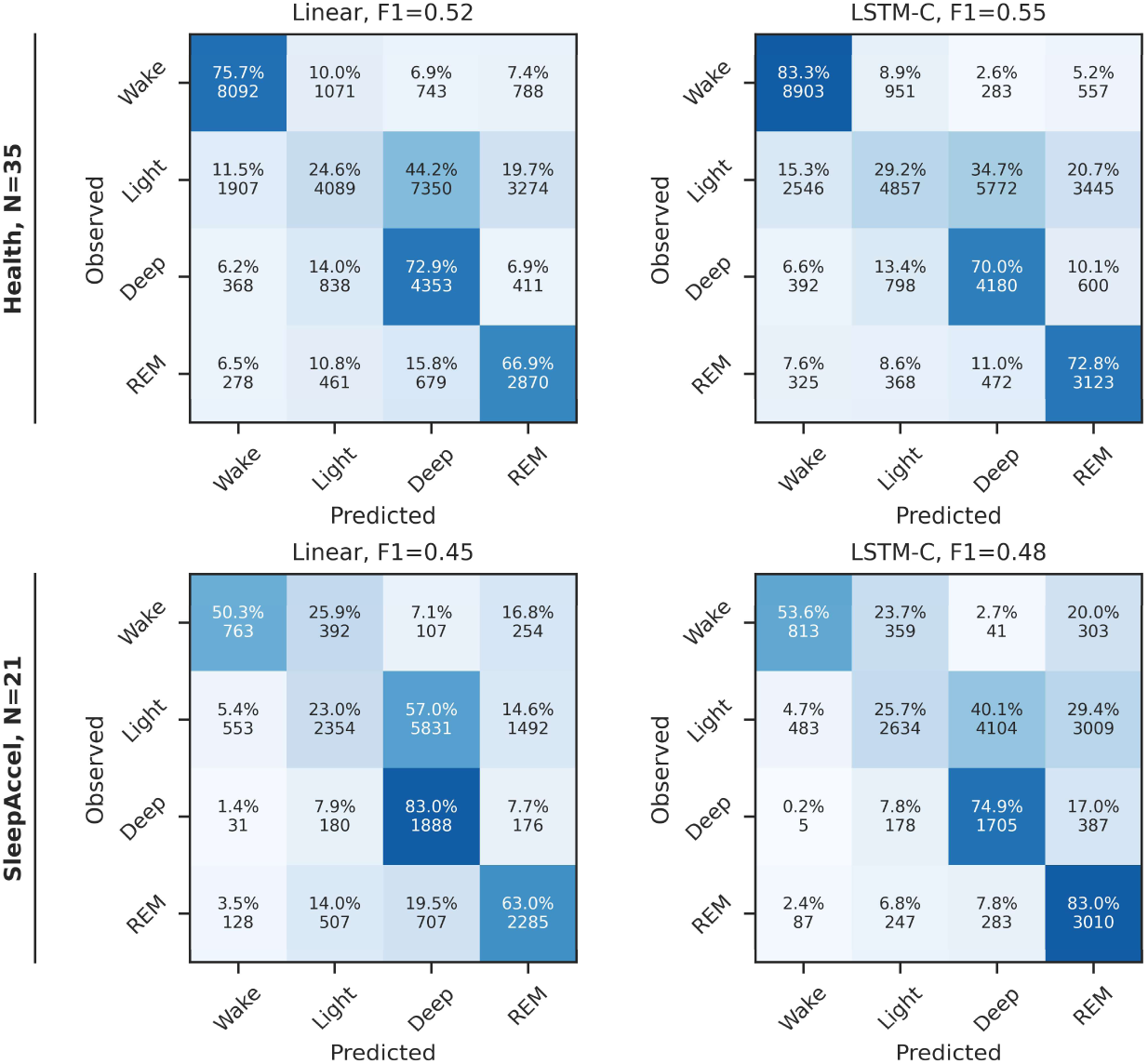
| Four-class sleep staging Confusion Matrices for external test sets.

**Supplementary Figure 22.**
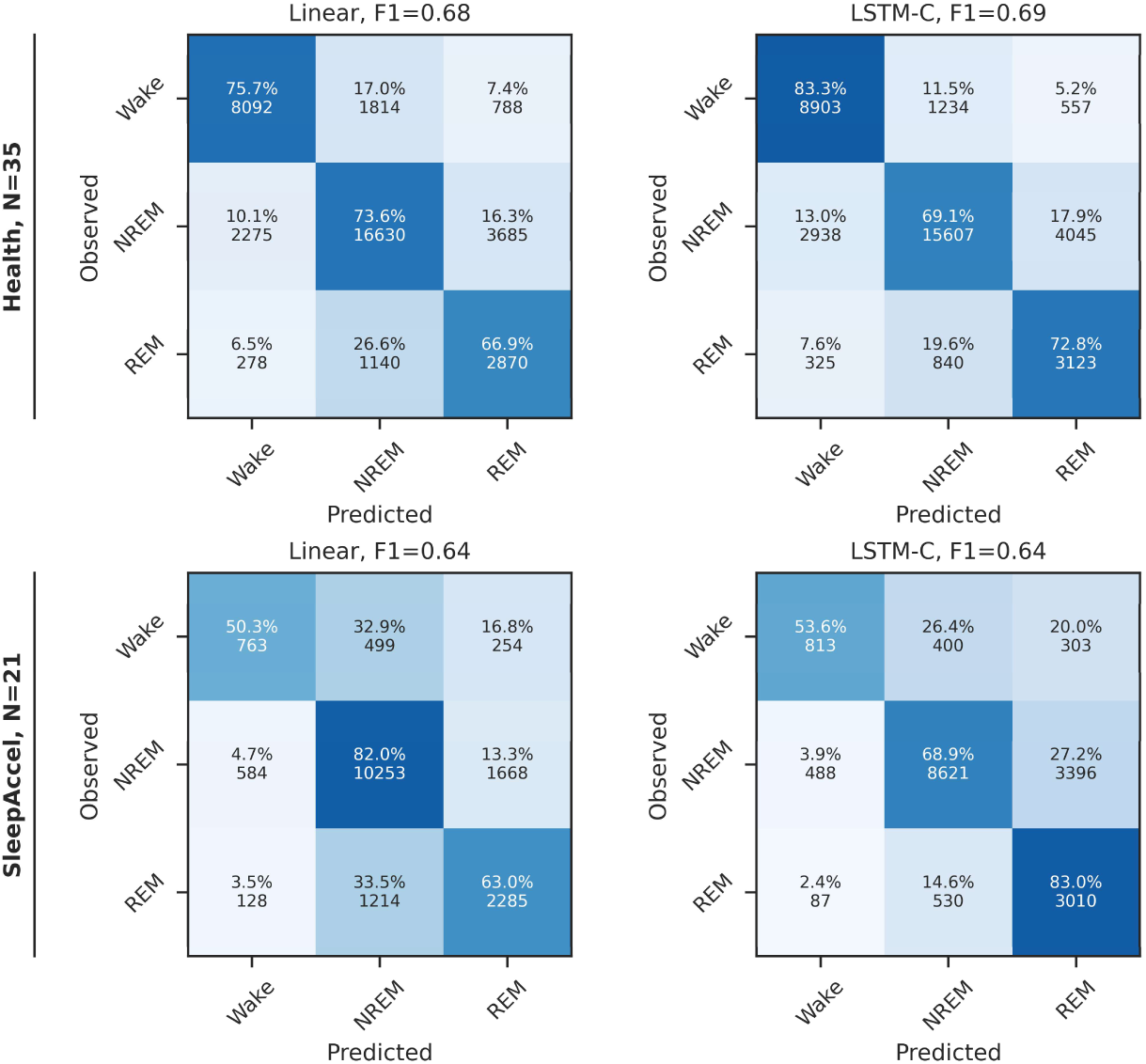
| Wake-NREM-REM confusion Matrices for external test sets.

**Supplementary Figure 23.**
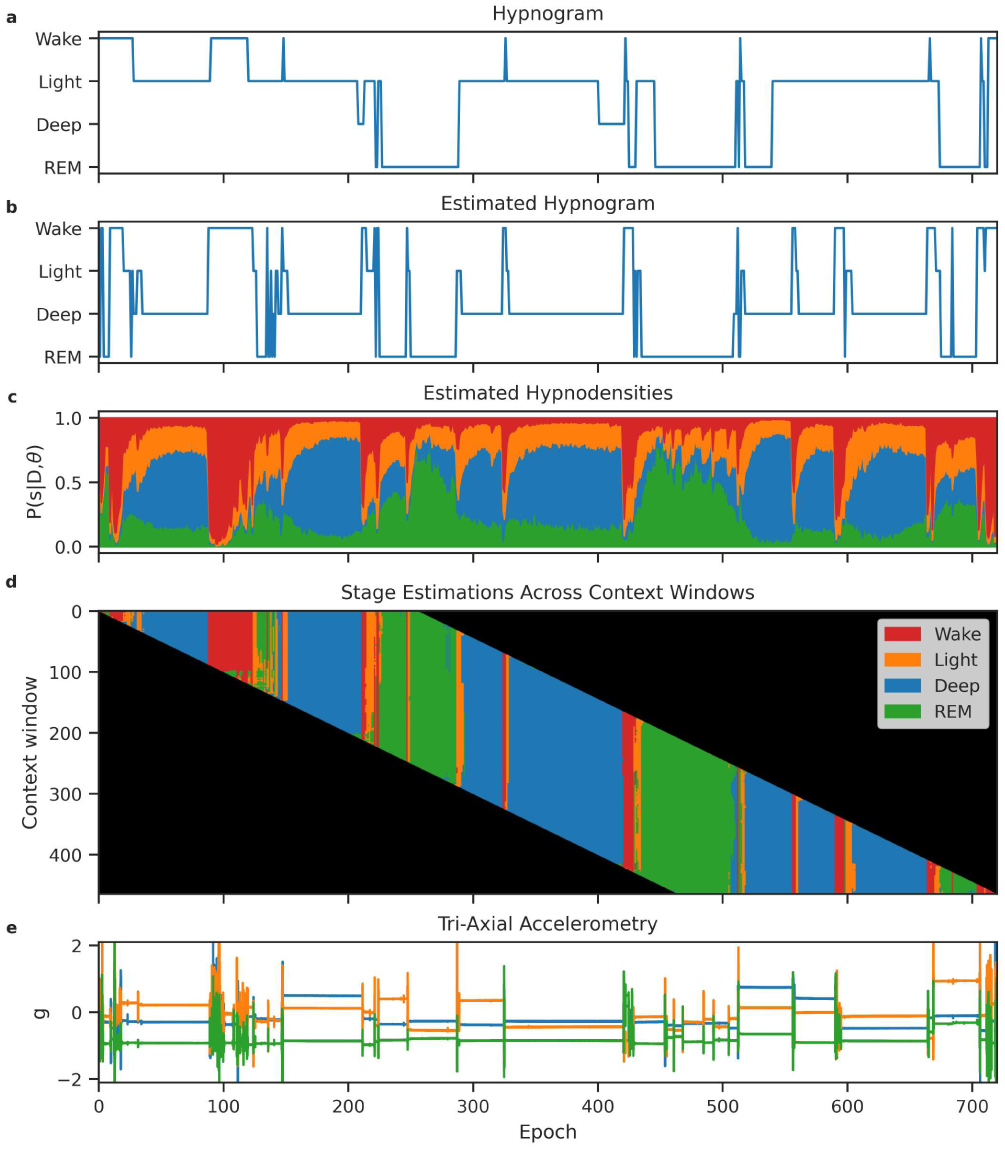
| AcceleRest Linear Sleep Stage Estimations for External Test Set Subject with Median Cohen’s Kappa. **a)** True hypnogram. **b)** estimated hypnogram. **c)** Estimated sleep stage probabilties (hypnodensities). **d)** Predicted sleep stages for each context window. **e)** Input tri-axial acceleormetry.

**Supplementary Figure 24.**
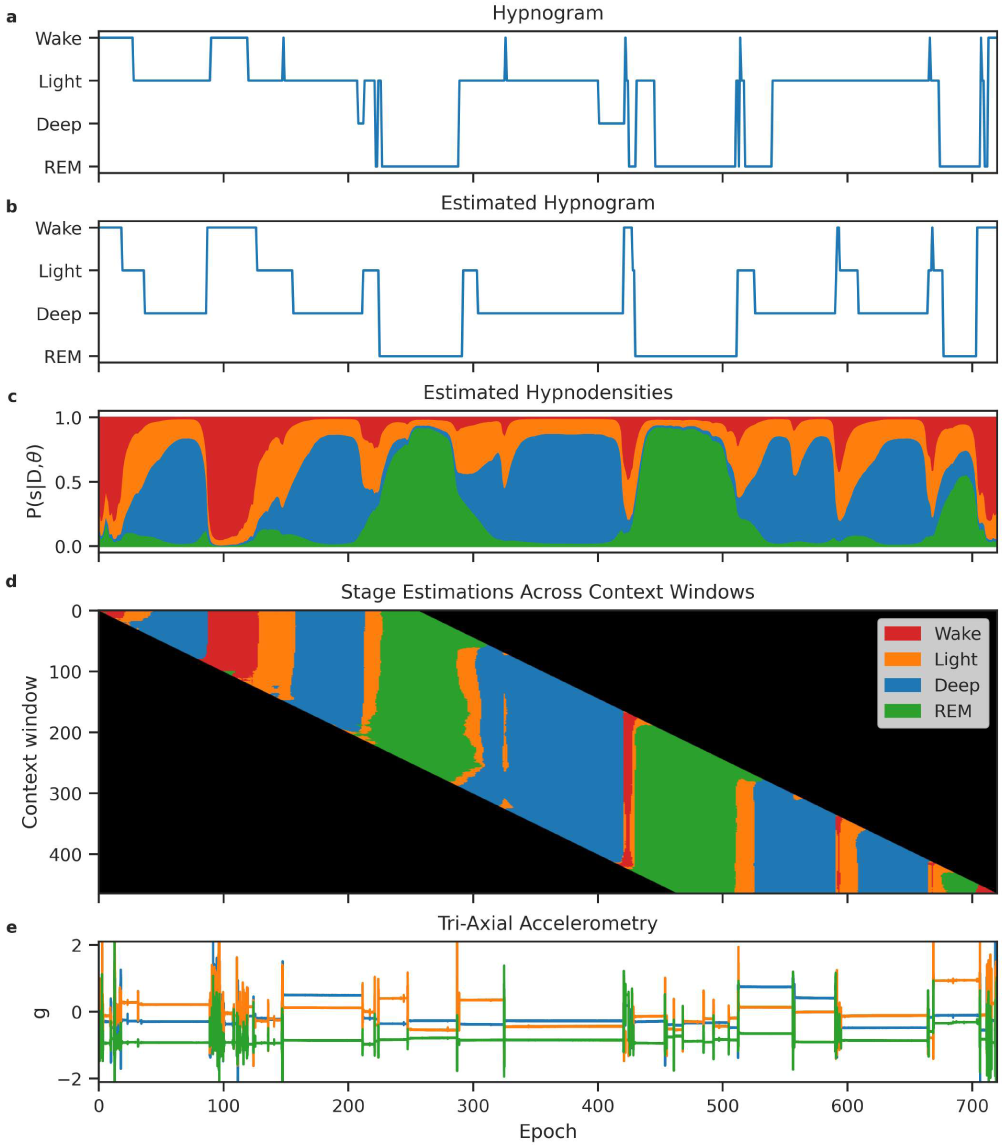
| AcceleRest LSTM-C Sleep Stage Estimations for External Test Set Subject with Median Cohen’s Kappa. **a)** True hypnogram. **b)** estimated hypnogram. **c)** Estimated sleep stage probabilties (hypnodensities). **d)** Predicted sleep stages for each context window. **e)** Input tri-axial acceleormetry.

**Supplementary Figure 25.**
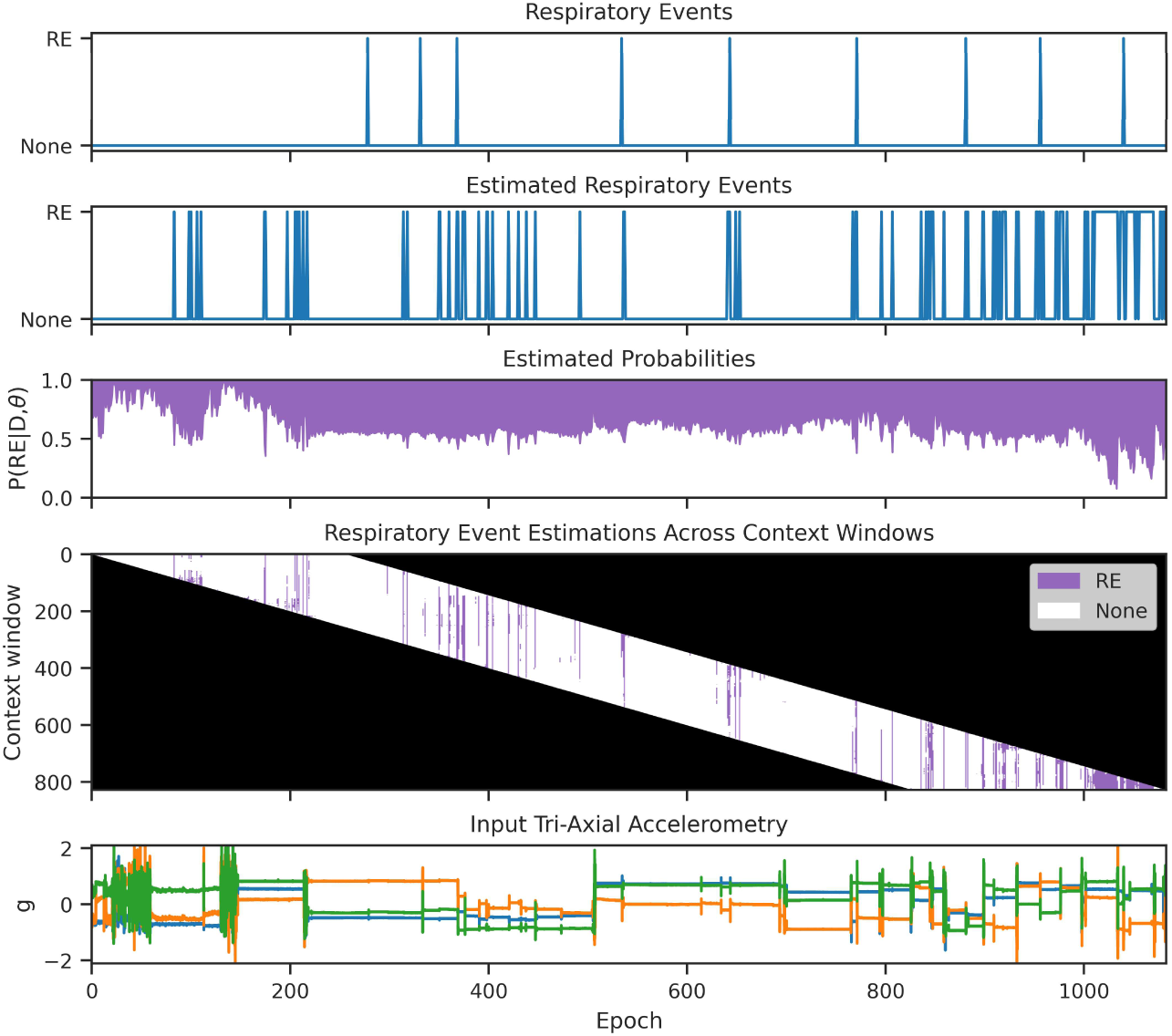
| Respiratory Event Predictions For External Test Subject with Median F1 Score. **a)** Annotated events. **b)** Predicted events. **c)** Estimated event probabilties. **d)** Predicted events for each context window. **e)** Input tri-axial acceleormetry.

**Supplementary Figure 26.**
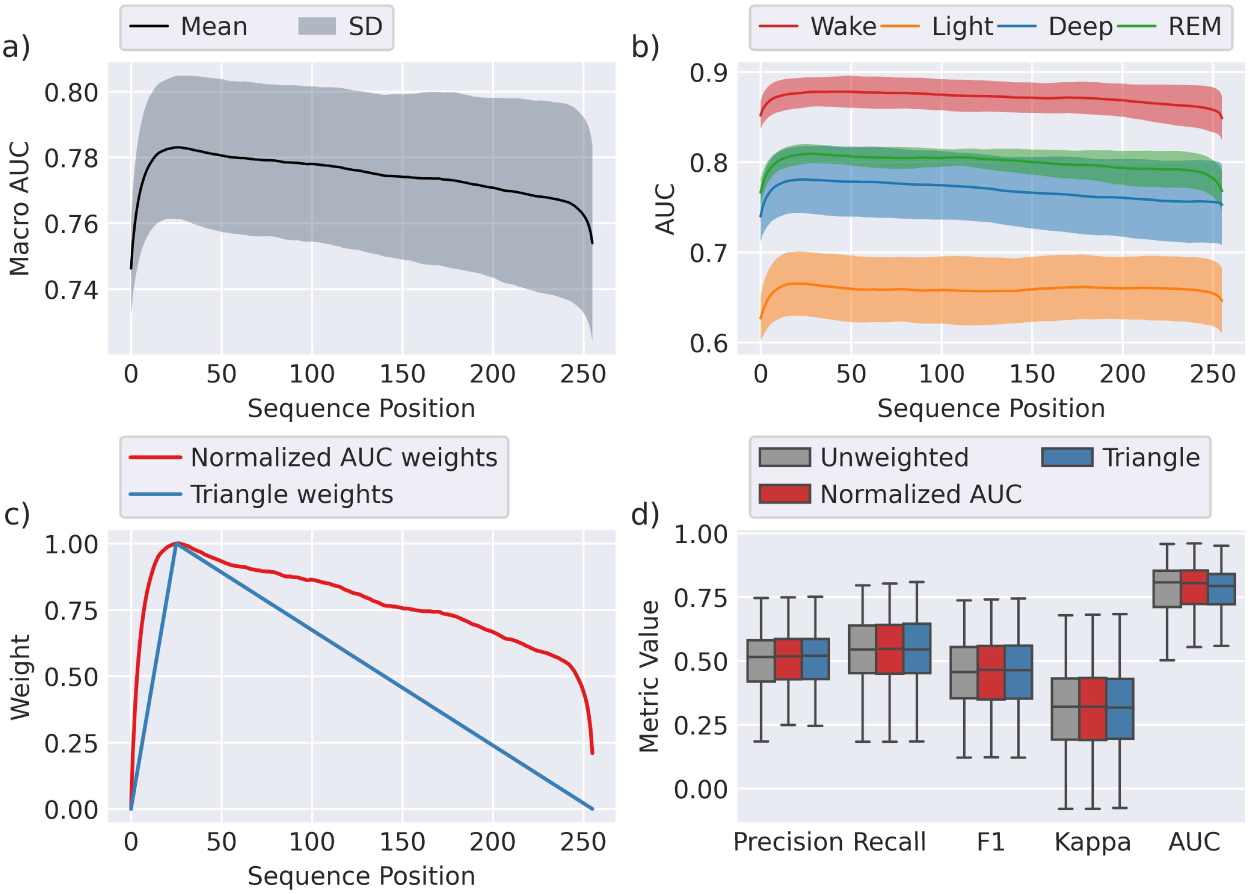
| AcceleRest performance across the context window. **a, b)** Macro and stage-wise area AUCs are area under the receiver operating characteristic (AUROC) for each position in the context window of AcceleRest, lines and fill show mean ± SD on 5-fold CV on the TBI training set. **c)** Sequence weights constructed from the normalized mean AUC or triangular weights. **d)** Subject-wise performances on the 5 validation folds with sliding-window predictions averaged with or without positional weights.

